# The impact of Mendelian sleep and circadian genetic variants in a population setting

**DOI:** 10.1101/2022.01.04.21268199

**Authors:** Michael N. Weedon, Samuel E. Jones, Jacqueline. M. Lane, Jiwon Lee, Hanna M. Ollila, Amy Dawes, Jess Tyrrell, Robin N. Beaumont, Timo Partonen, Ilona Merikanto, Stephen S. Rich, Jerome I. Rotter, Timothy M. Frayling, Martin K Rutter, Susan Redline, Tamar Sofer, Richa Saxena, Andrew R. Wood

## Abstract

Rare variants in ten genes have been reported to cause Mendelian sleep conditions characterised by extreme sleep duration or timing. These include familial natural short sleep (*ADRB1, DEC2/BHLHE41, GRM1 and NPSR1*), advanced sleep phase (*PER2, PER3, CRY2, CSNK1D* and *TIMELESS*) and delayed sleep phase (*CRY1*). The association of variants of these genes with extreme sleep conditions were usually based on clinically ascertained families, and their effects when identified in the population are unknown. We aimed to determine the effects of these variants on sleep traits in large population-based cohorts.

We performed genetic association analysis of variants previously reported to be causal for Mendelian sleep and circadian conditions. Analyses were performed using 191,929 individuals with data on sleep and whole-exome or genome-sequence data from 4 population-based studies: UK Biobank, FINRISK, Health-2000-2001, and the Multi-Ethnic Study of Atherosclerosis (MESA). We identified sleep disorders from self-report, hospital and primary care data. We estimated sleep duration and timing measures from self-report and accelerometery data.

We identified carriers for 10 out of 12 previously reported pathogenic variants for 8 of the 10 genes. They ranged in frequency from 1 individual with the variant in *CSNK1D* to 1,574 individuals with a reported variant in the *PER3* gene in the UK Biobank. We found no association of any of these variants with extreme sleep or circadian phenotypes. Using sleep timing as a proxy measure for sleep phase, only *PER3* and *CRY1* variants demonstrated association with earlier and later sleep timing, respectively; however, the magnitude of effect was smaller than previously reported (sleep midpoint ~7 mins earlier and ~5 mins later, respectively). We also performed burden tests of protein truncating (PTVs) or rare missense variants for the 10 genes. Only PTVs in *PER2* and *PER3* were associated with a relevant trait (for example, 64 individuals with a PTV in *PER2* had an odds ratio of 4.4 for being “definitely a morning person”, *P*=4×10^−8^; and had a 57-minute earlier midpoint sleep, *P*=5×10^−7^).

Our results indicate that previously reported variants for Mendelian sleep and circadian conditions are often not highly penetrant when ascertained incidentally from the general population.

## INTRODUCTION

Rare variants in ten genes have been reported to cause Mendelian sleep conditions that are characterised by extreme sleep duration or timing. For example, variants in the *ADRB1, NPSR1* and *GRM1* genes have been recently reported to cause familial natural short sleep among carriers, defined as 4 to 6 hours sleep with no adverse effects on mental health or well-being^1–3^. Short sleep duration has also been reported to be caused by variants in the *DEC2/BHLHE41* gene, a well-known Mendelian sleep gene^4^. Familial advanced sleep phase (FASP) where sleep timing is shifted 3 or 4 hours earlier has been reported for variants in *PER2*^5^, *PER3*^6^, *CRY2*^7^, *CSNK1D*^*8*^ and *TIMELESS*^*9*^. The opposite condition, familial delayed sleep phase disorder has been reported to be caused by a gain-of-function *CRY1* variant, c.1657+3A>C, with affected individuals sleeping approximately 1 hour later than unaffected individuals^10^.

The effect of variants in these genes on sleep duration and sleep timing when identified incidentally in the general population is unknown. Discovery efforts for these variants generally used either a single pedigree or a small number of families selected on a specific clinical phenotype. For example, for *ADRB1*, six individuals in a single family were affected with familial natural short sleep. This “phenotype first” method of discovery means we do not know the effect of these variants when identified in an individual from the general population (i.e. from a “genotype first” approach). It is important to re-evaluate effect estimates to understand the underlying biology which may inform clinical risk stratification, and because of the recent dramatic increase in direct-to-consumer (DTC) and health service genome-wide genetic testing. To assess the effect of these variants when identified incidentally, large, unselected population cohorts are needed.

Estimating the effects of these variants in the general population has not previously been possible due to limitations in the availability of genetic data coupled with sleep parameters. The UK Biobank, a population-based study of 500,000 individuals from the UK, provides an opportunity to address questions of pathogenicity and penetrance of rare genetic conditions^11^. We have previously shown, for a range of traits and diseases, that disease penetrance is generally lower in UK Biobank compared to that reported from clinical cohorts^12,13^. For example, using activity monitor derived and self-report estimates of sleep timing from the UK Biobank, we have demonstrated that the effect of the *PER3* P415A/H417R familial advanced sleep phase variant on sleep timing is substantially lower than the published estimate (0.13hrs vs. 4.2hrs)^14^. However, our previous studies were based on genotyping array data of relatively common single nucleotide polymorphisms (SNPs). Genotyping arrays are known to capture a relatively small number of rare coding pathogenic disease variants, typically with poor accuracy for genotyping and imputation^13,15,16^.

In this study, we use exome sequencing data in up to 184,065 individuals of European ancestry from the UK Biobank (October 2020 release) with sleep data, with additional data from up to 2,015 individuals from the Multi-Ethnic Study of Atherosclerosis (MESA), and 5,929 individuals from the FinnGen study to comprehensively assess the penetrance of Mendelian sleep and circadian genes in a population-based setting. We show that previously reported variants for Mendelian sleep and circadian conditions are often not highly penetrant when identified incidentally from the general population.

## METHODS

### UK Biobank Study Participants

The primary study population was drawn from the UK Biobank study – a longitudinal population-based study of individuals residing in the UK. We restricted our analysis to a subset of 184,532 Europeans with whole-exome sequence data, including 170,518 unrelated Europeans (<3^rd^ degree) defined through kinship coefficients made available from the UK Biobank. Details on derivation of genetic ancestry has previously been reported in Jones *et al*.^14^.

### Exome sequence data in UK Biobank

We used the second release of exome-sequence data from the UK Biobank (October 2020). Specifically, we use genotypes called and provided in binary PLINK format (data field: 23155). Genotypes for previously reported Mendelian causes of sleep and circadian conditions were extracted for subsequent data analysis. Details of central processing of whole-exome data on 200K UK Biobank participants can be found online as part of UK Biobank’s data showcase: https://biobank.ctsu.ox.ac.uk/crystal/label.cgi?id=170. All variants passed central quality control^17^. Sequence data for variants analysed in this paper were manually inspected through IGV^18^ plots.

### Sleep phenotypes and variant selection

We focussed our analysis on phenotypes previously reported to have Mendelian causes, specifically familial natural short sleep (≤6 hours)^1,3,4^, familial advanced sleep phase characterised by earlier sleep onset and earlier waking^5–9^, and delayed sleep phase disorder associated with later sleep onset and later waking^10^.

### Sleep disorders and medication

We used data from the UK Biobank variable 131061 (https://biobank.ndph.ox.ac.uk/showcase/field.cgi?id=131061) which classifies individuals as having a sleep disorder based on self-report, hospital and primary care data. As this variable does not separate out sub classifications of sleep disorders we used ICD-10 codes G472 and F512 from in-patient data to specifically assess disorders of the sleep wake cycle. Medication use at baseline in the UK Biobank was identified from variable 20003 (https://biobank.ctsu.ox.ac.uk/crystal/field.cgi?id=20003). Classification of sleep medication used in this paper has previously been described^19^. Since ‘sleep’ is a behavior, phenotyping is complicated. There are extreme sleep patterns like ASP and NSS which are often not considered ‘disorders’ by individuals if the trait does not interfere with an individual’s work and social demands^20^.

### Sleep Duration

We used self-reported sleep duration from UK Biobank questionnaire data (data field: 1160). We excluded individuals who reported >12 hours sleep duration, did not know or preferred not to answer. This data was also dichotomized for multiple analyses to define “short sleepers” as individuals self-reporting sleep duration ≤6 hours, ≤5 hours, ≤4 hours, and ≥4 hours ≤6. A maximum of 166,360 individuals had genotype and self-report sleep duration available across the variants previously reported to be Mendelian causes of familial natural short sleep and prioritised for analysis. In addition, we used accelerometer estimates of nocturnal sleep derived in a previous study^21^. A maximum subset of 34,241 individuals remained for analysis of accelerometer-based sleep duration estimates after removing individuals (n=4,323) with problematic accelerometer data processing or who were outliers for the number of nocturnal sleep episodes used to derive nocturnal sleep estimates^21^. There was no association for individual variants or overall with the individuals removed from the accelerometer analyses. We also applied additional quality control among non-carriers of previously reported variants described in this study by removing individuals with >12 hours estimated sleep duration.

### Sleep timing

We used self-reported diurnal preference available in the UK Biobank (data field: 1180) as a proxy for sleep timing, whereby we assumed morning people to sleep earlier and evening people to sleep later. We created four binary variables to represent diurnal preference where individuals were coded ‘1’ based on being: 1) definitely a morning person; 2) more or definitely a morning person; 3) definitely an evening person, and 4) more or definitely an evening person. For each variable, individuals who did not report having the respective circadian preference(s) (including “Do not know” but excluding “Prefer not to answer”) were coded ‘0’. A maximum of 168,409 individuals had genotype data and self-report chronotype data across the variants associated with advanced and delayed sleep phase. In addition, we used accelerometer estimates from up to 7 nights of the least-active 5 hours (L5) over a 24-hour period, with values representing hours from the previous midnight (e.g. 7□p.m.□=□19 and 2□a.m.□=□26)^21^. Sleep midpoint was estimated as the mid-point of the sleep period time window used to define sleep duration^21^. In total, a maximum subset of 34,650 individuals remained for statistical analysis of accelerometer-based sleep timing estimates. We applied additional quality control among non-carriers of previously reported variants described by removing individuals outside 4 standard deviations of the respective trait (midpoint sleep or L5 timing) analysed.

### Replication of Findings

To replicate our UK Biobank observations of previously reported variant-phenotype associations, we used self-reported measures of sleep duration and circadian preference (chronotype) in up to 5,929 individuals from two population-based studies from Finland: FINRISK^22^ and Health-2000-2011 (https://www.julkari.fi/handle/10024/130780), available through the FinnGen Project (https://www.finngen.fi/en). In addition, we used accelerometer-based estimates of sleep timing (sleep midpoint) in up to 1,935 individuals from the Multi-Ethnic Study of Atherosclerosis (MESA) Sleep Study^23^ where sequence-based genotypes of previously reported variants associated with sleep timing from its Exam 5 were available (see Supplementary Information for study descriptors).

### Statistical Analysis

#### Defining genotype groups for comparison of sleep parameter estimates

For each variant, summaries of sleep parameter estimates were analysed with by genotype. The set of individuals classified as homozygous reference for all variants was the same after removing individuals from this genotype group who were carriers for other variants.

#### Analysis of self-report phenotypes

t-tests tests were performed to compare means and standard deviations of the continuous self-report sleep duration variables across genotype groups and carrier status, respectively. For dichotomized variables, Fisher’s exact tests were performed to compare proportions of individuals labelled as short sleepers or had a defined circadian preference (above) across genotype groups. Alternate homozygous counts were combined with heterozygous counts when performing Fisher’s exact test where applicable. In addition, logistic regression was performed using UK Biobank data to obtain odds ratios, adjusting for age at baseline (field 21003), sex (field 31), assessment centre (field 54), month when attending the assessment centre (field 55), and 40 genetic principal components (field 22009) available from the UK Biobank.

#### Accelerometer-derived phenotypes

t-tests were performed to compare means and standard deviations of the accelerometer-based estimates of sleep duration, sleep-midpoint and L5 timing.

### Burden testing of rare loss-of-function and missense variants in UK Biobank

Genetic variants identified in the ten previously reported genes were annotated using the Ensembl Variant Effect Predictor (VEP)^24^ and LOFTEE^25^. Variants with MAF<0.0001 and annotated as missense or loss-of-function with high confidence were analysed through burden testing as implemented in Regenie^26^ that accounts for relatedness among individuals analysed. We analysed up to 184,065 individuals of inferred European genetic ancestry with available sleep and covariate data.

## RESULTS

### Most reported Mendelian sleep variants are present in UK Biobank and are not associated with sleep disorders

We assessed the frequencies of 12 variants in 10 genes that have been reported to cause familial natural short sleep, familial advanced sleep phase, or delayed sleep phase conditions (**Table 1**). All were present in the UK Biobank in unrelated individuals of European ancestry except for the S662G variant in *PER2* (not present in gnomAD (version 2.1)) and the Y206H variant in *NPSR1*. In addition, we identified carriers for previously reported variants in *GRM1, BHLHE41*/*DEC2, CRY1*, and *PER3* in the MESA and Finnish studies (**Table 2**). None of the variants were associated with any self-report or clinically diagnosed sleep disorder in UK Biobank, including the G472 or F512 ICD-10 code for disorders of the sleep wake cycle or with sleep medication use (**Supplementary Table 1**). We noted in the FinnGen study there was a nominal association with ICD10 code G472 (circadian rhythm sleep disorders) for the *CRY1* c.1657+3A>C variant (0.05% in controls vs. 0.29% in cases, *P*=0.026), but this variant is 10-fold rarer in the Finnish population than in non-Finnish Europeans and is imputed with a quality score of only 0.76.

**Table 1.** Summary of twelve variants previously reported to be causal for Mendelian sleep and circadian conditions, including the variant frequencies catalogued in gnomAD.

| PubMed ID | Gene | Variant | CHR:BP<sup>a</sup> | rsID<sup>b</sup> | Trait | gnomAD AAF<sup>f</sup> | gnomAD EAF<sup>g</sup> |
| --- | --- | --- | --- | --- | --- | --- | --- |
| 33065013 | *GRM1* | S458A | 6:146352435 | rs151255685 | FNSS<sup>c</sup> | $2 \times 10^{-4}$ | $2 \times 10^{-4}$ |
| 33065013 | *GRM1* | R889W | 6:146426563 | rs768023437 | FNSS<sup>c</sup> | $3 \times 10^{-5}$ | $6 \times 10^{-5}$ |
| 31619542 | *NPSR1* | Y206H | 7:34827538 | rs1406844918 | FNSS<sup>c</sup> | $4 \times 10^{-6}$ | $9 \times 10^{-6}$ |
| 31473062 | *ADRB1* | A187V | 10:114044692 | rs776439595 | FNSS<sup>c</sup> | $4 \times 10^{-5}$ | $8 \times 10^{-5}$ |
| 19679812 | *BHLHE41 (DEC2)* | P384R | 12:26122364 | rs121912617 | FNSS<sup>c</sup> | $3 \times 10^{-5}$ | $5 \times 10^{-5}$ |
| 28388406 | *CRY1* | c.1657+3A>C | 12:106992962 | rs184039278 | DSPD<sup>d</sup> | $4 \times 10^{-3}$ | $5 \times 10^{-3}$ |
| 26903630 | *PER3* | P415A | 1:7809893 | rs150812083 | FASP<sup>e</sup> | $6 \times 10^{-3}$ | $6 \times 10^{-3}$ |
| 26903630 | *PER3* | H417R | 1:7809900 | rs139315125 | FASP<sup>e</sup> | $6 \times 10^{-3}$ | $6 \times 10^{-3}$ |
| 11232563 | *PER2* | PER2S662G | 2:238257003 | rs121908635 | FASP<sup>e</sup> | - | - |
| 27529127 | *CRY2* | A260T | 11:45867648 | rs201220841 | FASP<sup>e</sup> | $6 \times 10^{-5}$ | $1 \times 10^{-4}$ |
| 31138685 | *TIMELESS* | R1081X | 12:56418347 | rs1465092391 | FASP<sup>e</sup> | $8 \times 10^{-6}$ | $2 \times 10^{-5}$ |
| 15800623 | *CSNK1D* | T44A | 17:82265743 | rs104894561 | FASP<sup>e</sup> | $4 \times 10^{-6}$ | $9 \times 10^{-6}$ |

<sup>a</sup>chromosome and base-pair position defined by the Genome Reference Consortium Human Build 38; <sup>b</sup>reference SNP (rs) cluster identifier of variant; <sup>c</sup>familial natural short sleep; <sup>d</sup>delayed sleep phase disorder; <sup>e</sup>familial advanced sleep phase; <sup>f</sup>allele frequency of minor allele across all ancestries catalogued in gnomAD (version 2.1). <sup>g</sup>allele frequency of minor allele within samples of European ancestry (excluding Finnish) catalogued in gnomAD (version 2.1).

**Table 2.** Maximum genotype counts for 12 previously reported monogenic causes of sleep and circadian conditions in unrelated individuals of European ancestry from the UK Biobank, FinnGen and MESA studies. Genotype counts are based on availability of sleep characteristics relevant to each gene.

| Gene / Variant | Trait | REF/ALT<sup>d</sup> | UK Biobank |  |  | FINNGEN |  |  | MESA |  |  |
| --- | --- | --- | --- | --- | --- | --- | --- | --- | --- | --- | --- |
|  |  |  | REF/REF<sup>e</sup> | REF/ALT<sup>f</sup> | ALT/ALT<sup>g</sup> | REF/REF<sup>e</sup> | REF/ALT<sup>f</sup> | ALT/ALT<sup>g</sup> | REF/REF<sup>e</sup> | REF/ALT<sup>f</sup> | ALT/ALT<sup>g</sup> |
| *GRM1* / S458A | FNSS<sup>a</sup> | T/G | 169,451 | 67 | - | 5,927 | 2 | 0 | - | - | - |
| *GRM1* / R889W | FNSS<sup>a</sup> | A/T | 169,513 | 3 | - | - | - | - | - | - | - |
| *NPSR1* / Y206H | FNSS<sup>a</sup> | T/C | - | - | - | - | - | - | - | - | - |
| *ADRB1* / A187V | FNSS<sup>a</sup> | C/T | 169,450 | 69 | - | - | - | - | - | - | - |
| *BHLHE41 (DEC2)* / P384R | FNSS<sup>a</sup> | G/C | 169,500 | 10 | - | - | - | - | 1,993 | 22 | - |
| *CRY1* / c.1657+3A>C | DSPD<sup>b</sup> | T/G | 168,586 | 1,480 | 9 | 2,838 | 4 | 0 | - | - | - |
| *PER3* / P415A | FASP<sup>c</sup> | C/G | 168,500 | 1,565 | 7 | 2,734 | 149 | 3 | 2,003 | 12 | - |
| *PER3* / H417R | FASP<sup>c</sup> | A/G | 168,500 | 1,567 | 7 | 2,734 | 149 | 3 | 2,003 | 12 | - |
| *PER2* / PER2S662G | FASP<sup>c</sup> | A/G | - | - | - | - | - | - | - | - | - |
| *CRY2* / A260T | FASP<sup>c</sup> | G/A | 170,038 | 38 | - | - | - | - | - | - | - |
| *TIMELESS* / R1081X | FASP<sup>c</sup> | G/A | 170,064 | 5 | - | - | - | - | - | - | - |
| *CSNK1D* / T44A | FASP<sup>c</sup> | T/C | 170,075 | 1 | - | - | - | - | - | - | - |

<sup>a</sup>familial natural short sleep; <sup>b</sup>delayed sleep phase disorder; <sup>c</sup>familial advanced sleep phase; <sup>d</sup>reference allele / alternate allele; <sup>e</sup>number of homozygous carriers for reference allele; <sup>f</sup>number of heterozygous carriers; <sup>g</sup>number of homozygous carriers for alternate allele.

### ADRB1, GRM1, or DEC2/BHLHE41 pathogenic variants are not associated with self-reported short sleep duration in population-based cohorts

We identified 149 unrelated individuals from the UK Biobank with self-reported measures of sleep duration and carrying a previously reported pathogenic variant for natural short sleep in *ADRB1 (*A817V, *n*=69*), DEC2/BHLHE41 (*P384R, *n*=10), or *GRM1 (*S458A, *n*=67; A889T, *n*=3). We found no evidence that these individuals have short sleep durations in the UK Biobank (**Table 3** and **Supplementary Tables 2** and **3**). For example, the 69 carriers of the *ADRB1* variant had a self-reported average sleep duration of 7.1 hours (95% CI: 6.9, 7.3) compared to 7.2 hours (95% CI: 7.19, 7.21) among non-variant carriers (t-test *P*=0.62). The proportion of *ADRB1* variant carriers self-reporting ≤6 hours of sleep, 23.2%, was no different to the proportion of people not carrying the variant, 23.7% (Fisher’s exact *P*=1.00). Using a fully adjusted logistic regression model gave similar results with an odds ratio of 0.98 (95% CI: 0.56–1.72; *P*=0.96) for sleeping less than 6 hours. We observed zero carriers of the *ADRB1* variant self-reporting a more extreme phenotype of ≤4 hours of sleep. Similar observations were made for previously reported monogenic sleep disruption variants in *DEC2/BHLHE41* and *GRM1*. The 10 carriers of the P384R variant in the *DEC2/BHLHE41* gene had an average self-reported sleep duration of 7.3 hours compared to 7.2 hours among non-carriers (*P*=0.69), with 10% of carriers reporting ≤6 hours sleep duration compared to 23.7% among non-carriers of reported variants (P=0.47). Carriers for variants in the *GRM1* gene did not significantly differ in sleep duration from non-carriers (S458A: 7.1 hours (carriers) vs 7.2 hours (non-carriers), P=0.81; A889T: 8 hours (carriers) vs 7.2 hours (non-carriers), P=0.18), or the number of individuals reporting sleep duration of ≤6 hours (S458A: 28.4% (carriers) vs 23.7% (non-carriers), P=0.39; A889T: 0% (carriers) vs 23.8% (non-carriers), P=1.00).The null effect of the S458A variant in *GRM1* was also observed in FinnGen where 2 S458A carriers were identified and had no statistically significant difference compared to non-carriers in average sleep duration (t-test *P*=0.92) or proportion self-reporting ≤6 hours sleep (Fisher’s exact *P*=1.00) (**Table 3**).

**Table 3.** Summary of self-reported sleep duration in UK Biobank (data field 1160) between carriers of variants previously reported to be causal for familial natural short sleep and non-carriers (homozygous reference called by UK Biobank exome-sequencing) who are also non-carriers for any of the other 12 variants described in this article.

| | | | | | | Self-report sleep duration (hrs) | | | | Reporting $\leq 6$ hours sleep | | |
| --- | --- | --- | --- | --- | --- | --- | --- | --- | --- | --- | --- | --- |
| Gene | Variant | REF/ALT<sup>a</sup> | Study | Genotype | N<sup>b</sup> | Min<sup>c</sup> | Max<sup>d</sup> | Mean (SD<sup>e</sup>) | P<sup>f</sup> | %<sup>g</sup> | Cases / Controls | P<sup>h</sup> |
| *ADRB1* | A187V | C/T | UKB | C/C | 166,291 | 1 | 12 | 7.17 (1.07) | 0.62 | 23.74 | 39,484 / 126,807 | 1.00 |
|  |  |  |  | C/T | 69 | 5 | 12 | 7.10 (1.03) |  | 23.19 | 16 / 53 |  |
| *BHLHE41* | P384R | G/C | UKB | G/G | 166,283 | 1 | 12 | 7.17 (1.07) | 0.69 | 23.74 | 39,483 / 126,800 | 0.47 |
|  |  |  |  | G/C | 10 | 5 | 9 | 7.30 (1.06) |  | 10.00 | 1 / 9 |  |
| *GRM1* | S458A | T/G | UKB | T/T | 166,290 | 1 | 12 | 7.17 (1.07) | 0.81 | 23.74 | 39,484 / 126,806 | 0.39 |
|  |  |  |  | T/G | 67 | 4 | 10 | 7.13 (1.15) |  | 28.36 | 19 / 48 |  |
|  |  |  | FinnGen | T/T | 5,927 | 3 | 15 | 7.59 (1.20) | 0.92 | 14.10 | 837 / 5,090 | 1.00 |
|  |  |  |  | T/G | 2 | 7 | 8 | 7.50 (0.71) |  | 0.00 | 0 / 2 |  |
|  | A889T | A/T | UKB | A/A | 166,288 | 1 | 12 | 7.17 (1.07) | 0.18 | 23.75 | 39,485 / 126,803 | 1.00 |
|  |  |  |  | A/T | 3 | 8 | 8 | 8.00 (0.00) |  | 0.00 | 0 / 3 |  |

<sup>a</sup>reference allele / alternate allele; <sup>b</sup>number of individuals in genotype group; <sup>c</sup>minimum sleep duration self-reported in genotype group;
<sup>d</sup>maximum sleep duration self-reported in genotype group; <sup>e</sup>standard deviation; <sup>f</sup>t-test *P*-value (two-sided); <sup>g</sup>%=percentage of individuals in genotype group self-reporting $\leq 6$ hours sleep duration (cases); <sup>h</sup>Fisher's exact-test *P*-value (two-sided).

### ADBR1, GRM1, or DEC2/BHLHE41 pathogenic variants are not associated with accelerometer derived measures of sleep in population based cohorts

We confirmed the lack of association between previously identified pathogenic variants and sleep duration using accelerometer estimates of sleep in a subset of 34,226 individuals from the UK Biobank. Fifteen *ADRB1* variant carriers with accelerometer-derived sleep estimates had an average sleep duration of 7.6 hours (95% CI: 7.4, 7.8) compared to 7.3 hours (95% CI: 7.29, 7.31) among non-variant carriers (t-test P=0.20) (**Table 4**). All 15 *ADRB1* variant carriers had an accelerometer-based sleep duration average of more than 6 hours (min=6 hours, 53 minutes). Similar observations were made when stratifying accelerometer data analyses to weekend nights and weekday nights (**Supplementary Table 4**).

**Table 4.** Summary of average accelerometer derived sleep duration (hours) (all nights) in the subset of exome-sequenced unrelated Europeans in UK Biobank, split by for carriers and non-carriers of variants previously reported to be causal for familial natural short sleep.

| Gene | Variant | Genotype | N<sup>a</sup> | Min<sup>b</sup> | Max<sup>c</sup> | Mean | SD<sup>d</sup> | P<sup>e</sup> |
| --- | --- | --- | --- | --- | --- | --- | --- | --- |
| *ADRB1* | A187V | C/C | 34,168 | 1.63 | 11.87 | 7.30 | 0.86 | 0.20 |
|  |  | C/T | 15 | 6.89 | 8.83 | 7.59 | 0.49 |  |
| *BHLHE4 (DEC2)* | P384R | G/G | 34,167 | 1.63 | 11.87 | 7.30 | 0.86 | 0.62 |
|  |  | G/C | 4 | 6.26 | 8.25 | 7.51 | 0.92 |  |
| *GRM1* | S458A | T/T | 34,166 | 1.63 | 11.87 | 7.30 | 0.86 | 0.92 |
|  |  | T/G | 10 | 6.48 | 8.30 | 7.33 | 0.76 |  |
|  | A889T | A/A | 34,167 | 1.63 | 11.87 | 7.30 | 0.86 | - |
|  |  | A/T | 0 | - | - | - | - |  |

<sup>a</sup>number of individuals in genotype group; <sup>b</sup>minimum average sleep duration in genotype group; <sup>c</sup>maximum average sleep duration in genotype group; <sup>d</sup>standard deviation; <sup>e</sup>t-test *P*-value (two-sided).
<sup>a</sup>percentage of individuals in genotype group self-reporting being “definitely a ‘morning’ person”; <sup>b</sup>Fisher’s exact-test *P*-value (two-sided); <sup>c</sup>number of individuals; <sup>d</sup>minimum phenotypic value; <sup>e</sup>maximum phenotypic value; <sup>f</sup>SD=standard deviation; <sup>g</sup>carrier status t-test *P*-value (two-sided). Homozygous carriers for rare alleles combined with heterozygote carriers when performing Fisher’s exact-test.

### PER3, but not CRY2 or TIMELESS, variants are associated with advanced sleep phase in the population-based cohorts, but with reduced effect size

Variants in five genes have previously been associated with familial advanced sleep phase syndrome – characterised by approximately ≥3 hour shifts towards earlier sleep and wake times. We previously tested the *PER3* P415A/H417R variant in the UK Biobank and found it was associated with self-report diurnal preference and activity monitor derived sleep timing, although the size of the effect on sleep timing (L5 time) was smaller than the initially published estimate of 4.2 hours (7.8 minutes, 95% CI: 4.2–13.2 minutes, *P*=4.3×10^−4^). We confirmed this association using exome sequence data and accelerometer data. The difference on average sleep-midpoint timing between carriers and non-carriers based on exome-sequence data was 6.8 minutes (95% CI: 1.4-12.3 minutes, *P*=0.01) with a similar effect size for L5 timing (**Table 5**). Variant carriers had an odds ratio of 1.36 (95% CI: 1.22 – 1.52, P=2×10^−8^) for “definitely” being a morning person. We found no evidence that carriers of previously reported pathogenic variants in the other two genes, *CRY2* and *TIMELESS*, had altered chronotype, L5 timing or sleep-midpoint indicative of earlier sleep timing (**Supplementary Tables 5-7**).

**Table 5.**
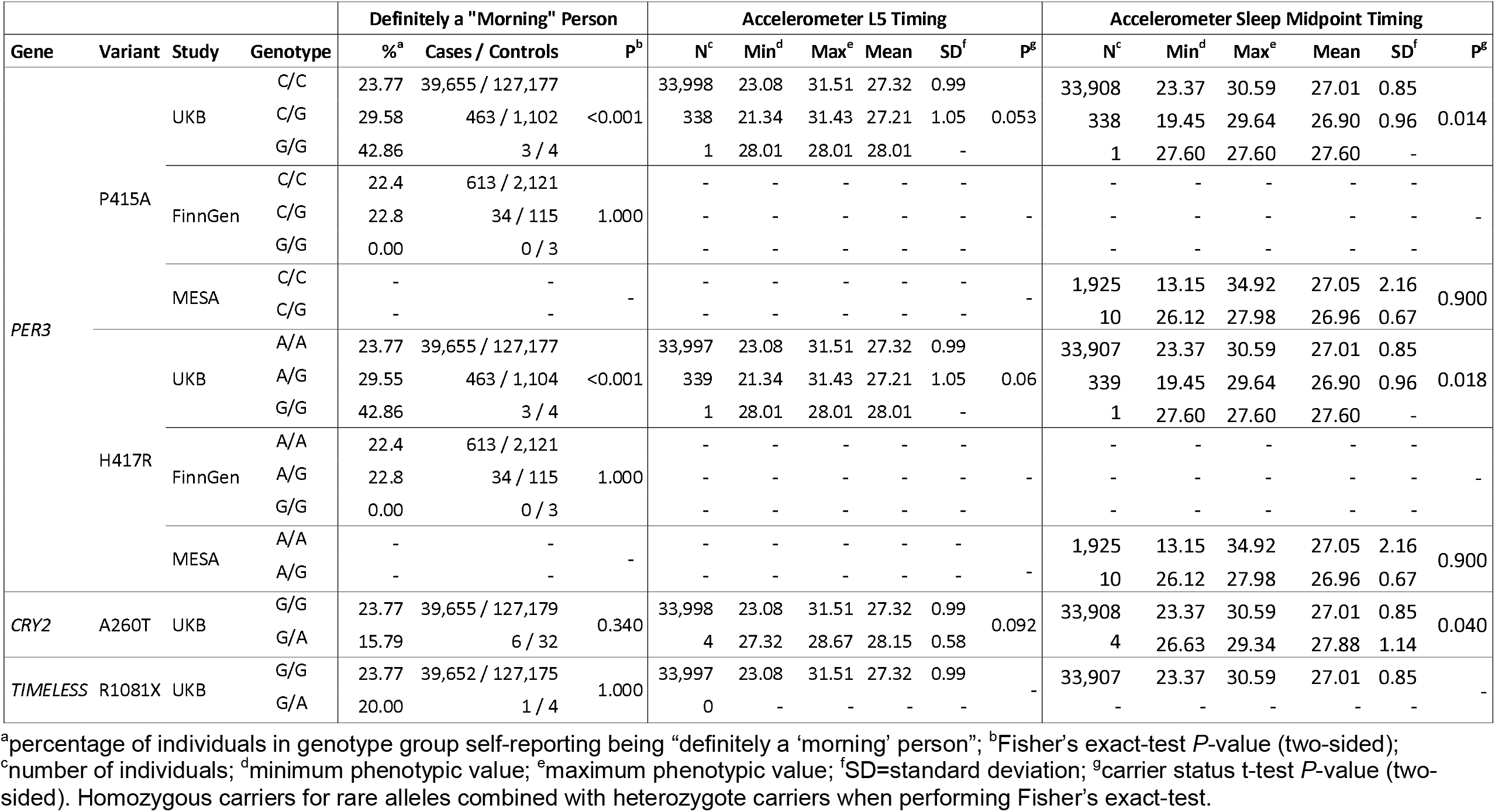
Proportion of individuals self-reporting as being “definitely a morning person”, average L5 timing and average of sleep-midpoint across all nights for each genotype group of variants previously reported to be causal for familial advanced sleep phase in the UK Biobank (UKB), Finnish and MESA studies. Accelerometer-based estimates of sleep timing unavailable in the Finnish studies. Self-reported “Morningness” and accelerometer estimates of L5-timing unavailable in MESA.

### *CRY1* c.1657+3A>C *is associated with diurnal preference and a delayed sleep phase in a population-based cohort, but with reduced effect size*

*CRY1* c.1657+3A>C has previously been associated with delayed sleep phase disorder, characterised by an approximately 1-hour shift towards later sleep and wake times. In the UK Biobank, 10.1% and 11.1% of *CRY1* heterozygous and homozygous variant carriers, respectively, reported being “definitely an evening person”, compared to 7.9 % of non-variant carriers (Fisher’s exact *P*=0.003) (**Table 6**). No carriers of the *CRY1* variant reported being “definitely an evening person” in the Finnish studies. The observed difference in the UK Biobank in sleep-midpoint estimated from accelerometery for individuals with a *CRY1* variant was 5.4 minutes later (95% CI:-0.2,11.0, *P*=0.06), with a similar difference observed for L5 timing. Similar point estimates were observed in our sensitivity analyses that included restricting accelerometer data analyses to either weekend nights or weekday nights (**Supplementary Tables 8-10** and **Supplementary Figs 1-3**).

**Table 6.** Proportion of individuals self-reporting as being “definitely an evening person”, average L5 timing, and average sleep-midpoint across all nights for each genotype group of the *CRY1* variant previously reported to be causal for delayed sleep phase disorder in the UK Biobank (UKB), Finnish and MESA studies. Accelerometer based estimates of sleep timing unavailable in the Finnish studies. Self-reported “Eveningness” and accelerometer estimates of L5-timing unavailable in MESA.

| Gene | Variant | Study | Genotype | Definitely an "Evening" Person |  |  | Accelerometer L5 Timing |  |  |  |  |  | Accelerometer Sleep Midpoint |  |  |  |  |  |
| --- | --- | --- | --- | --- | --- | --- | --- | --- | --- | --- | --- | --- | --- | --- | --- | --- | --- | --- |
|  |  |  |  | %<sup>a</sup> | Cases/Controls | P<sup>b</sup> | N<sup>c</sup> | Min<sup>d</sup> | Max<sup>e</sup> | Mean | SD<sup>f</sup> | P<sup>g</sup> | N<sup>c</sup> | Min<sup>d</sup> | Max<sup>e</sup> | Mean | SD<sup>f</sup> | P<sup>g</sup> |
| CRY1 | c.1657+3A>C | UKB | T/T | 7.92 | 13,212 / 153,621 |  | 33,998 | 23.08 | 31.51 | 27.32 | 0.99 |  | 33,908 | 23.37 | 30.59 | 27.01 | 0.85 |  |
|  |  |  | T/G | 10.07 | 149 / 1,331 | 0.003 | 318 | 24.06 | 30.38 | 27.41 | 1.00 | 0.132 | 318 | 24.28 | 29.60 | 27.11 | 0.88 | 0.059 |
|  |  |  | G/G | 11.11 | 1 / 8 |  | 4 | 25.08 | 28.44 | 26.50 | 1.48 |  | 4 | 25.49 | 27.38 | 26.50 | 0.90 |  |
|  |  | FinnGen | T/T | 10 | 284 / 2,554 | 1.000 | - | - | - | - | - | - | - | - | - | - | - | - |
|  |  |  | T/G | 0 | 0 / 4 |  | - | - | - | - | - | - | - | - | - | - | - | - |
|  |  | MESA | T/T | - | - | - | - | - | - | - | - | - | 1,914 | 13.19 | 34.92 | 27.05 | 2.14 | 0.568 |
|  |  |  | T/G | - | - |  | - | - | - | - | - | - | 21 | 13.15 | 30.10 | 26.78 | 3.32 |  |

<sup>a</sup>percentage of individuals in genotype group self-reporting being “definitely a ‘evening person””; <sup>b</sup>Fisher’s exact-test *P*-value (two-sided); <sup>c</sup>number of individuals.; <sup>d</sup>minimum phenotypic value; <sup>e</sup>maximum phenotypic value; <sup>f</sup>standard deviation; <sup>g</sup>carrier status t-test *P*-value (two-sided). Homozygous carriers for rare alleles combined with heterozygote carriers when performing Fisher’s exact-test.

### Heterozygous protein truncating variants in reported Mendelian sleep genes PER2 and PER3 are associated with sleep timing

Most of the reported variants are missense variants and some, for example, *CRY1* c.1657+3A>C have been shown to have a specific gain of function effect in *in-vitro* experiments^10^. Only the reported *TIMELESS* gene variant is a protein truncating variant (PTV) and it is unclear whether the reported Mendelian sleep variants act through loss of function due to haploinsufficiency. We therefore identified all rare (MAF<0.01%) nonsense, frameshift and essential splice site variants across the ten Mendelian sleep and circadian genes. The number of individuals with rare high confidence loss-of-function variants in these genes ranged from 10 for *ADRB1* to 205 for *TIMELESS*. We subsequently performed burden testing of loss-of-function variants for these 10 genes in up to 184,065 individuals of European ancestry, including and adjusting for relatedness (**Supplementary Tables 11-14**). We identified associations between *PER2* and self-reported measures of diurnal preference and accelerometer-estimates of sleep timing. We observed associations between loss-of-function variants in the *PER2* gene and UK Biobank participants self-reporting as “definitely a morning person” (Burden *P*=4×10^−8^) (**Supplementary Table 12**), and accelerometer-estimates of sleep-midpoint (Burden *P*=5×10^−7^) and L5 timing (Burden *P*=9×10^−4^)(**Supplementary Table 14**). Of 64 unrelated European carriers carrying at least one of the 50 loss-of-function variants in the *PER2* gene, 58% (n=37) self-reported as “definitely a morning person” in contrast to 24% (n=40,438) among non-carriers (Fisher’s exact *P*<0.0001) (**Supplementary Table 15**). Within unrelated individuals, compared with non-carriers of loss-of-function variants in *PER2*, carriers had an earlier average sleep-midpoint of ~57 minutes (t-test *P*<0.0001) (**Supplementary Table 16)** and an earlier average L5 timing of ~33 minutes (t-test *P*=0.027) (**Supplementary Table 17**). In addition, our burden testing identified an association between loss-of-function variants in *PER3* and L5-timing (Burden *P*=4×10^−6^) (**Supplementary Table 14**). Our gene-based analyses for high confidence PTVs did not result in associations for other previously reported monogenic genes for sleep duration or timing. We observed no associations in gene-based tests of rare missense variants in these genes after accounting for multiple testing (Bonferroni *P*=0.0001 based on 42 tests across 10 genes).

## DISCUSSION

Recent studies have identified 10 genes where specific variants are reported to cause familial natural short sleep, familial advanced sleep phase, or delayed sleep phase. These studies have tended to be based on a limited number of families ascertained to have a specific sleep trait. This form of ascertainment means the effect of the variants and genes when identified in the population is unknown. Here, we show that most previously reported variants for Mendelian sleep and circadian conditions are not highly penetrant when ascertained incidentally from the general population. Incidental findings are becoming increasingly common with the increase in whole genome sequencing both from direct-to-consumer companies and through health services. It is important, therefore, to get accurate estimates of the risk of developing a condition so that individuals are not misdiagnosed and/or potentially incorrectly treated for a condition.

We and others have shown previously^13^ that the penetrance of rare Mendelian disease variants is likely to be lower when estimated from population-based cohorts than in ascertained discovery or clinical cohorts and this may be the case for the genes and variants reported here. It is also possible that some of the reported genes and variants are not causes of the reported sleep conditions in humans. The functional effect of each the variants assessed here are supported by extensive *in vitro* and animal model studies. The level of human genetic evidence varies across studies, from 6 individuals from a single pedigree for the *ADBR1* variant to 78 individuals from 7 families for the CRY1 c.1657+3A>C variant. It is therefore possible that some of the reported variants with weaker human genetic evidence do not cause monogenic sleep and circadian conditions in humans. However, because of the nature of the ascertainment, our study cannot address pathogenicity and can only conclude that the effect of these variants when identified incidentally from the population appears to be much weaker than previously reported.

There are several other possible explanations for the differences between our studies. Genetic background may play a role. Sleep is so important to survival, that homeostatic mechanisms are extremely robust. For example, the very strong circadian phenotype of homozygous *PER2* KO allele^27^ was completely absent when crossed onto a C57/Bl6 background^28^. Our work underscores the significant challenges of behavioral genetics It is possible that an individual or family presenting to a clinic with a specific sleep or circadian condition have an increased polygenic susceptibility to sleep duration, in addition to the monogenic variant. For example, we have recently identified 351 variants for being a morning person from genome-wide association studies. Individuals in the highest 5% of polygenic risk had an average sleep timing of 25 mins earlier compared to the lowest polygenic risk individuals. As has been shown for traits such as lipids and BMI^29^, this suggests that extreme polygenic risk can have similar effect sizes to monogenic variants for sleep traits. Ancestry differences may also play a role. For example, the *CRY1* c.1657+3A>C association was discovered in an American family and followed up in Turkish families. A recent paper has found association with the *CRY1* variants and sleep-midpoint timing (~40 mins) in an independent cohort of Turkish ancestry individuals^30^. The difference in effect size between these studies may be due to different genetic backgrounds between these previous populations and the predominantly Northern European ancestry population used in our analyses. There are also potential environmental (e.g. daylight hours) and societal explanations for the different results in this study compared to previous studies.

There are several limitations to our study which provide other possible explanations for the weaker associations observed here compared to previous studies. First, the UK Biobank has a healthy volunteer bias^31^ and may select against individuals with sleep disorders. However, this is unlikely for FASP and FNSS where the phenotypes of phase advance or short sleep rarely affect individuals’ well-being^20^. Additionally, the allele frequency of the variants in UK Biobank is similar to that in a large resource of exome data (gnomAD) suggesting limited selection against these variants. Second, reported sleep patterns that are often shaped by social factors, and thus may not reflect their underlying sleep preferences. This could explain the lack of association with sleep timing for example. However, we find no association with the variants with traits such as self-reported ease of getting up and limiting the analyses to an individual’s activity on the weekend shows a similar lack of association. Additionally, we find no association with any measures of sleep quality or disruption in the UK Biobank, including those defined by medical record codings, although sleep fragmentation has been observed in *CRY1* c.1657+3A>C carriers^10^.

Another limitation to this study is that it is not possible to do as detailed sleep and circadian phenotyping in this large-scale study as is possible in smaller scale clinical studies. We have, however, used multiple data sources including self-report and activity monitor data and have used primary care and inpatient data to identify sleep disorders. We have previously validated the activity monitor sleep estimates against polysomnography data^32^ and it is a reliable measure of many sleep parameters. The validity of the measures is also confirmed by the statistically robust associations with *PER2* and *PER3* protein truncating variants using both self-report and accelerometry, with effects of sleeping timing of approximately 1 hour. We have also demonstrated this through the robust association of hundreds of common genetic variants with chronotype^14^, sleep duration^33^ and other sleep measures through genome-wide association studies (GWAS)^19,21^. The number of individuals with variants is relatively low for some genes, but the number of individuals carrying previously identified variants is usually larger than the number of available in the original reports.

Our work demonstrates that haploinsufficiency of *PER2* affects circadian timing in humans. We find a substantial effect on chronotype and sleep timing for individuals with heterozygous *PER2* protein truncating variants. This is unexpected because only homozygous *Per2* knockout mice exhibit a circadian shortened circadian period, with no phenotype in heterozygotes^34^. The human genetic evidence for a role of the *PER2* S662G missense variant is robust with co-segregation in a large pedigree with FASPS^5^. It was initially thought that the effect of this S662G variant was caused by decreased phosphorylation of PER by CK1ε that could stabilize it leading to PER accumulating prematurely and shortened circadian period. Others have suggested that the S662G mutation results in decreased PER2 levels and/or an increased turnover of nuclear PER2^35^. Our work shows that, in humans, haploinsufficiency of *PER2* causes a substantial effect on diurnal preference and sleeping timing.

Our results indicate that most previously reported variants for Mendelian sleep and circadian conditions are not highly penetrant causes of extreme sleep duration or timing when ascertained incidentally from the population.

## Data Availability

Data is available upon application to the relevant studies.

## ACKNOWLEDGEMENTS

We thank Louis Ptáček, Alina Patke and Michael Young for helpful discussion and comments on the manuscript. R.N.B is supported by grant MR/T00200X/1. Whole genome sequencing (WGS) for the Trans-Omics in Precision Medicine (TOPMed) program was supported by the National Heart, Lung and Blood Institute (NHLBI). WGS for “NHLBI TOPMed: Multi-Ethnic Study of Atherosclerosis (MESA)” (phs001416.v1.p1) was performed at the Broad Institute of MIT and Harvard (3U54HG003067-13S1). Core support including centralized genomic read mapping and genotype calling, along with variant quality metrics and filtering were provided by the TOPMed Informatics Research Center (3R01HL-117626-02S1; contract HHSN268201800002I). Core support including phenotype harmonization, data management, sample-identity QC, and general program coordination were provided by the TOPMed Data Coordinating Center (R01HL-120393; U01HL-120393; contract HHSN268201800001I). We gratefully acknowledge the studies and participants who provided biological samples and data for MESA and TOPMed. JSK was supported by the Pulmonary Fibrosis Foundation Scholars Award and grant K23-HL-150301 from the NHLBI. MRA was supported by grant K23-HL-150280, AJP was supported by grant K23-HL-140199, and AM was supported by R01-HL131565 from the NHLBI. EJB was supported by grant K23-AR-075112 from the National Institute of Arthritis and Musculoskeletal and Skin Diseases.

The MESA project is conducted and supported by the National Heart, Lung, and Blood Institute (NHLBI) in collaboration with MESA investigators. Support for MESA is provided by contracts 75N92020D00001, HHSN268201500003I, N01-HC-95159, 75N92020D00005, N01-HC-95160, 75N92020D00002, N01-HC-95161, 75N92020D00003, N01-HC-95162, 75N92020D00006, N01-HC-95163, 75N92020D00004, N01-HC-95164, 75N92020D00007, N01-HC-95165, N01-HC-95166, N01-HC-95167, N01-HC-95168, N01-HC-95169, UL1-TR-000040, UL1-TR-001079, and UL1-TR-001420. Also supported in part by the National Center for Advancing Translational Sciences, CTSI grant UL1TR001881, and the National Institute of Diabetes and Digestive and Kidney Disease Diabetes Research Center (DRC) grant DK063491 to the Southern California Diabetes Endocrinology Research Center.

## ETHICS AND CONSENT

The UK Biobank was granted ethical approval by the North West Multi-centre Research Ethics Committee (MREC) to collect and distribute data and samples from the participants (http://www.ukbiobank.ac.uk/ethics/) and covers the work in this study, which was performed under UK Biobank application numbers 9072 and 16434. All participants included in these analyses gave informed consent to participate.

## DATA AVAILABILITY

The datasets analyzed for this study are available on application to the participating studies.

## REFERENCES

1. Shi G, Xing L, Wu D, et al. A Rare Mutation of beta1-Adrenergic Receptor Affects Sleep/Wake Behaviors. Neuron 2019; 103(6): 1044–55 e7.

2. Shi G, Yin C, Fan Z, et al. Mutations in Metabotropic Glutamate Receptor 1 Contribute to Natural Short Sleep Trait. Curr Biol 2020.

3. Xing L, Shi G, Mostovoy Y, et al. Mutant neuropeptide S receptor reduces sleep duration with preserved memory consolidation. Sci Transl Med 2019; 11(514).

4. He Y, Jones CR, Fujiki N, et al. The transcriptional repressor DEC2 regulates sleep length in mammals. Science 2009; 325(5942): 866–70.

5. Toh KL, Jones CR, He Y, et al. An hPer2 phosphorylation site mutation in familial advanced sleep phase syndrome. Science 2001; 291(5506): 1040–3.

6. Zhang L, Hirano A, Hsu PK, et al. A PERIOD3 variant causes a circadian phenotype and is associated with a seasonal mood trait. Proc Natl Acad Sci U S A 2016; 113(11): E1536–44.

7. Hirano A, Shi G, Jones CR, et al. A Cryptochrome 2 mutation yields advanced sleep phase in humans. Elife 2016; 5.

8. Xu Y, Padiath QS, Shapiro RE, et al. Functional consequences of a CKIdelta mutation causing familial advanced sleep phase syndrome. Nature 2005; 434(7033): 640–4.

9. Kurien P, Hsu PK, Leon J, et al. TIMELESS mutation alters phase responsiveness and causes advanced sleep phase. Proc Natl Acad Sci U S A 2019; 116(24): 12045–53.

10. Patke A, Murphy PJ, Onat OE, et al. Mutation of the Human Circadian Clock Gene CRY1 in Familial Delayed Sleep Phase Disorder. Cell 2017; 169(2): 203–15 e13.

11. Sudlow C, Gallacher J, Allen N, et al. UK biobank: an open access resource for identifying the causes of a wide range of complex diseases of middle and old age. PLoS Med 2015; 12(3): e1001779.

12. Tuke MA, Ruth KS, Wood AR, et al. Mosaic Turner syndrome shows reduced penetrance in an adult population study. Genet Med 2019; 21(4): 877–86.

13. Wright CF, West B, Tuke M, et al. Assessing the Pathogenicity, Penetrance, and Expressivity of Putative Disease-Causing Variants in a Population Setting. Am J Hum Genet 2019; 104(2): 275–86.

14. Jones SE, Lane JM, Wood AR, et al. Genome-wide association analyses of chronotype in 697,828 individuals provides insights into circadian rhythms. Nat Commun 2019; 10(1): 343.

15. Weedon MN, Jackson L, Harrison JW, et al. Use of SNP chips to detect rare pathogenic variants: retrospective, population based diagnostic evaluation. BMJ 2021; 372: n214.

16. Weedon MN, Jackson L, Harrison JW, et al. Assessing the analytical validity of SNP-chips for detecting very rare pathogenic variants: implications for direct-to-consumer genetic testing. bioRxiv, doi: 101101/696799 2019.

17. Szustakowski JD, Balasubramanian S, Kvikstad E, et al. Advancing human genetics research and drug discovery through exome sequencing of the UK Biobank. Nat Genet 2021; 53(7): 942–8.

18. Robinson JT, Thorvaldsdottir H, Winckler W, et al. Integrative genomics viewer. Nat Biotechnol 2011; 29(1): 24–6.

19. Lane JM, Jones SE, Dashti HS, et al. Biological and clinical insights from genetics of insomnia symptoms. Nat Genet 2019; 51(3): 387–93.

20. Ashbrook LH, Krystal AD, Fu YH, Ptacek LJ. Genetics of the human circadian clock and sleep homeostat. Neuropsychopharmacology 2020; 45(1): 45–54.

21. Pulit SL, Stoneman C, Morris AP, et al. Meta-analysis of genome-wide association studies for body fat distribution in 694 649 individuals of European ancestry. Hum Mol Genet 2019; 28(1): 166–74.

22. Borodulin K, Tolonen H, Jousilahti P, et al. Cohort Profile: The National FINRISK Study. Int J Epidemiol 2018; 47(3): 696–i.

23. Bild DE, Bluemke DA, Burke GL, et al. Multi-Ethnic Study of Atherosclerosis: objectives and design. Am J Epidemiol 2002; 156(9): 871–81.

24. McLaren W, Gil L, Hunt SE, et al. The Ensembl Variant Effect Predictor. Genome Biol 2016; 17(1): 122.

25. Karczewski KJ, Francioli LC, Tiao G, et al. The mutational constraint spectrum quantified from variation in 141,456 humans. Nature 2020; 581(7809): 434–43.

26. Mbatchou J, Barnard L, Backman J, et al. Computationally efficient whole genome regression for quantitative and binary traits. BioRxiv 2021.

27. Bae K, Jin X, Maywood ES, Hastings MH, Reppert SM, Weaver DR. Differential functions of mPer1, mPer2, and mPer3 in the SCN circadian clock. Neuron 2001; 30(2): 525–36.

28. Xu Y, Toh KL, Jones CR, Shin JY, Fu YH, Ptacek LJ. Modeling of a human circadian mutation yields insights into clock regulation by PER2. Cell 2007; 128(1): 59–70.

29. Khera AV, Chaffin M, Aragam KG, et al. Genome-wide polygenic scores for common diseases identify individuals with risk equivalent to monogenic mutations. Nat Genet 2018; 50(9): 1219–24.

30. Smieszek SP, Brzezynski JL, Kaden AR, et al. An Observational Study Investigating the CRY1Δ11 Variant Associated with Delayed Sleep-wake Patterns and Circadian Metabolic Output. PREPRINT Research Square, DOI: 1021203/rs3rs-468649/v1 2021.

31. Munafo MR, Tilling K, Taylor AE, Evans DM, Davey Smith G. Collider scope: when selection bias can substantially influence observed associations. Int J Epidemiol 2018; 47(1): 226–35.

32. van Hees VT, Sabia S, Jones SE, et al. Estimating sleep parameters using an accelerometer without sleep diary. Sci Rep 2018; 8(1): 12975.

33. Dashti HS, Jones SE, Wood AR, et al. Genome-wide association study identifies genetic loci for self-reported habitual sleep duration supported by accelerometer-derived estimates. Nat Commun 2019; 10(1): 1100.

34. Zheng B, Larkin DW, Albrecht U, et al. The mPer2 gene encodes a functional component of the mammalian circadian clock. Nature 1999; 400(6740): 169–73.

35. Takahashi JS, Hong HK, Ko CH, McDearmon EL. The genetics of mammalian circadian order and disorder: implications for physiology and disease. Nat Rev Genet 2008; 9(10): 764–75.

