## Supplementary Methods for "The impact of Mendelian sleep and circadian genetic variants in a population setting"

**Description of Additional Studies**

***Multi-Ethnic Study of Atherosclerosis (MESA)***

The MESA study is a longitudinal study of risk factors for subclinical and clinical cardiovascular disease (CVD)^1^ begun in 2000-2002, with recruitment of 6,814 men and women aged 45-84, free of known CVD, from 6 communities across the U.S. Sampling aimed to achieve a racially/ethnically diverse sample, and included individuals who were non-Hispanic white (38%), black (28%), Hispanic (22%), and Asian (12%, predominantly Chinese descent). Subsequent examinations occurred to collect follow-up information and additional detained phenotyping. In conjunction with its fifth examination (2010-2013), a sleep ancillary study was conducted, which included 7-day actigraphy, in-home polysomnography, and a sleep questionnaire. All MESA participants provided written informed consent, and the study was approved by the Institutional Review Boards at The Lundquist Institute (formerly Los Angeles BioMedical Research Institute) at Harbor-UCLA Medical Center, University of Washington, Wake Forest School of Medicine, Northwestern University, University of Minnesota, Columbia University, and Johns Hopkins University.

*Sleep phenotypes.* Objective sleep-wake data was available for 2,261 individuals, collected using 7-day wrist actigraphs (Actiwatch Spectrum; Philips Respironics, Murrysville, PA), as described before^2^. Average sleep-midpoint timing was estimated for all nights, weekday nights, and weekend nights. For quality control, we required that sleep duration, either self-reported or derived by actigraphy, would be at least 1 hour and no higher than 20 hours, and excluded individuals with values outside this range. Sleep timing variables were centered at midnight, and individuals with sleep timing ≥11 hours before or after midnight were excluded.

*Whole genome sequencing.* Whole Genome Sequencing in MESA was performed via the Trans-Omics for Precision Medicine (TOPMed) program. WGS was performed using DNA from blood at the Broad Institute of MIT and Harvard using Illumina X10 technology at an average sequencing depth of >30X. We used the MESA call set generated as part of TOPMed Freeze 8 release. Details regarding sequencing methods and quality control are provided elsewhere^3^ (<https://www.nhlbiwgs.org/data-sets>).

***FinnGen***

Two Finnish cohorts were available with both exome-sequence data and sleep measures recorded in the same individuals: FINRISK and Health 2000-2011.

*FINRISK.* The FINRISK study was a Finnish population-based study into non-communicable disease interventions initially targeting cholesterol, blood pressure and smoking prevalence but later expanded to cover further outcomes. Recruitment initially began in 1972 with new waves of recruitment every five years. After the latest recruitment wave in 2012, over 100,000 individuals, aged 25 to 75 years, had participated. Participants were invited to a health exam and/or to complete a health- and lifestyle-based questionnaire. Amongst the questions asked were those on sleep habits, though only insomnia-related questions were asked prior to 2007. Further details on the FINRISK cohort are provided elsewhere^4^.

*FINRISK sleep phenotypes.* Self-report habitual sleep information and exome-sequence data were only available in the same individuals from the FINRISK 2007 wave so only these individuals from FINRISK were included in any analysis. For assessing habitual sleep duration, we used the answer to the question “How many hours on average do you sleep a) in one night? and b) in a period of 24 hours?” to which the participant was asked to report to the nearest hour and could freely enter the number. For a fair comparison with the UK Biobank self-report sleep duration, we used the answer to the latter question (“in a period of 24 hours?”). To assess diurnal preference, we used the answer to the question *There are so-called “morning people” (early to rise, early to bed) and “evening people” (late to rise, late to bed). Which are you?* to which the participants could select one of *absolutely a “morning person”*, *more “morning” than “evening person”*, *more “evening” than “morning person”* or *absolutely an “evening person”*. As with the UK Biobank data, we characterised people as either ‘morning people’ (those selecting *absolutely a “morning person”* or *more “morning” than “evening person”*) or ‘evening people’ (those selecting *more “evening” than “morning person”* or *absolutely an “evening person”*). Questions relating to habitual workday and free-day bedtimes and wake times were asked but only in the 2012 wave and so these measures were not used due to lack of sequence data in the 2012 recruitment wave. Additional information about the FINRISK questionnaires can be found at <https://thl.fi/en/web/thlfi-en/research-and-development/research-and-projects/the-national-finrisk-study/questionnaires>

*FINRISK whole-exome sequencing.* Venous blood samples were taken from participants at the health exam, separated by centrifuge at the health exam sites and (for the FINRISK 2007 and 2012 waves) stored locally then transported to the central THL laboratory twice a week. DNA extracted from these blood samples was exome-sequenced to a high-depth at three different facilities.Approximately 88% of the samples (with both phenotype and sequence data) were sequenced at the Wellcome Trust Sanger Institute (Hinxton, UK) on the Illumina HiSeq 2000 platform with a Roche SeqCap EZ HGSC VCRome exome reagent, generating 2x100bp paired-end sequence data with a minimum coverage depth of 20x for at least 70% of targeted bases in each sample. Around 8% of the samples included in analyses were exome-sequenced at the McDonnell Genome Institute (Washington University, USA) with a Roche SeqCap EZ HGSC VCRome reagent and performed with either the Illumina HiSeq 2000 (2x-100bp paired-end reads) or 2500 1T (2x125bp paired end reads) sequencer. Samples with a minimum coverage of 20x for at least 70% of targeted (exonic) bases were included variant calling and subsequent analyses. The mean coverage across all samples was 87x. The remaining 4% of included samples were sequenced at the Broad Institute (Cambridge, MA, USA) on an Illumina HiSeq 2000 or 2500 1T platform with a Roche SeqCap EZ HGSC VCRome exome reagent^5^.

*Health-2000-2011.* Health 2000 and Health 2011 (referred to collectively as Health-2000-2011) were Finnish population-based studies focussed on understanding the healthcare needs of Finns based on demographic and geographic factors. The Health 2000 survey collected data on 10,000 individuals who were 18 years of age or over and were sampled randomly from the population. The Health 2011 survey collected additional information on 8,135 participants recruited in the Health 2000 survey, 1,994 new participants aged 18-28 and on a further 920 participants of the unrelated Mini-Finland Survey. Participants were invited for a detailed health examination where anthropometric measures and blood samples were taken. At these examinations, participants were asked to complete an extensive questionnaire with information on demographic, lifestyle and health factors. Details on participants who did not attend the health examination were collected either through a phone interview or using questionnaires send by post. Several questions pertaining to sleep habits were recorded in both surveys. Further methodological details are available elsewhere (<https://www.julkari.fi/handle/10024/130780>).

*Health-2000-2011 sleep phenotypes.* To assess sleep duration, participants were asked in Health 2000 to answer the question *How many hours do you sleep in 24 hours?* and in Health 2011 the equivalent question *How many hours on average do you sleep in 24 hours?* and were asked to answer each to the nearest hour. With individuals who provided valid responses in both 2000 and 2011, we used the response from the 2000 survey. No measures related to circadian preference or habitual sleep onset and wake times were recorded in this cohort. Details on available measures and the exact questions used are available at <https://thl.fi/en/web/thl-biobank/for-researchers/sample-collections/health-2000-and-2011-surveys>.

*Health-2000-2011 exome sequencing.* Participants of Health-2000-2011 provided multiple venous blood samples at the health examination, one of which was retained and frozen at -20°C within 90 minutes of collection. Samples were transported and stored at the laboratories of the National Public Health Institute of Finland (KTL) until the were sent for whole-exome and whole-genome sequencing at the Broad Institute (Cambridge, MA, USA) and the Wellcome Trust Sanger Institute (Hinxton, UK). Of the samples included in the analyses, 92% were whole-exome sequenced at the Broad Institute, 7% were exome-sequenced at the Wellcome Trust Sanger Institute.

*FINRISK and Health-2000-2011 sequence alignment and variant calling.* Alignment and variant calling for both FINRISK and H2000 samples were performed together at the Broad Institute (Cambridge, MA, USA). Sequence data was aligned to genome reference build 37 using bwa-mem v0.7.7, then indels were realigned using GATK^6^, duplicate reads marked with picard (“Picard Toolkit.” 2019. Broad Institute, GitHub Repository, <http://broadinstitute.github.io/picard/>; Broad Institute) and clipping of overlapping bases using bamUtil v1.0.11^7^. Samples were then excluded if SNV calls were less than 90% concordant with existing genotype array data on the same samples, if the estimated contamination was over 3% or if there was evidence of a sample swap. Variants were excluded if they failed to pass GATK’s VSQR 99% filter (99.9% filter for WGS samples), had >2% missingness, had a Hardy-Weinberg P-value of <10^-6^ or with an overall allele balance of <30% in genotyped samples. Finally, samples were then excluded if they had >2% missing variant calls. More details on sequencing and quality control of these samples can be found elsewhere^5,8^.

*Monogenic sleep variants called in FINRISK and Health-2000-2011.* Of the four variants associated with FNSS, rs151255685 and rs121912617 were present in FINRISK and only rs151255685 was available in Health-2000-2011. The DSPS variant rs184039278, was present in both FINRISK and Health-2000-2011. Two of the six FASP variants, rs150812083 and rs139315125, were present in both FINRISK and Health-2000-2011. When considering individuals with a non-missing genotype for at least one of these five variants and at least one valid sleep measure, there were 2,915 samples from FINRISK and 3,759 from Health-2000-2011 (total N=6,674).

**REFERENCES**

1.	Bild DE, Bluemke DA, Burke GL, et al. Multi-Ethnic Study of Atherosclerosis: objectives and design. *Am J Epidemiol* 2002; **156**(9): 871-81.

2.	Ogilvie RP, Redline S, Bertoni AG, et al. Actigraphy Measured Sleep Indices and Adiposity: The Multi-Ethnic Study of Atherosclerosis (MESA). *Sleep* 2016; **39**(9): 1701-8.

3.	Taliun D, Harris DN, Kessler MD, et al. Sequencing of 53,831 diverse genomes from the NHLBI TOPMed Program. *Nature* 2021; **590**(7845): 290-9.

4.	Borodulin K, Tolonen H, Jousilahti P, et al. Cohort Profile: The National FINRISK Study. *Int J Epidemiol* 2018; **47**(3): 696-i.

5.	Ganna A, Satterstrom FK, Zekavat SM, et al. Quantifying the Impact of Rare and Ultra-rare Coding Variation across the Phenotypic Spectrum. *Am J Hum Genet* 2018; **102**(6): 1204-11.

6.	DePristo MA, Banks E, Poplin R, et al. A framework for variation discovery and genotyping using next-generation DNA sequencing data. *Nat Genet* 2011; **43**(5): 491-8.

7.	Jun G, Wing MK, Abecasis GR, Kang HM. An efficient and scalable analysis framework for variant extraction and refinement from population-scale DNA sequence data. *Genome Res* 2015; **25**(6): 918-25.

8.	Locke AE, Steinberg KM, Chiang CWK, et al. Exome sequencing of Finnish isolates enhances rare-variant association power. *Nature* 2019; **572**(7769): 323-8.
