## Supplementary Tables for "The impact of Mendelian sleep and circadian genetic variants in a population setting"

Supplementary Table 1. Sleep disorder and medication status by carrier status of variants previously reported to affect sleep duration or timing. Sleep disorders in the UK Biobank coded as G47 from self-report, ICD-10 codes or primary care data. Also presented are results for disorders of the sleep wake cycle (ICD10: G472). Sleep medication codes reported at UK Biobank baseline at reported in Lane *et al.* Nature Genetics, 2019.

|  |  |  | **Any self-report, primary care or ICD-10**  **code sleep disorder records (G47)** | | | | **Any sleep medication** | | | | **Any ICD-10 code for sleep-wake**  **disorder (G472 or F512)** | | | |
| --- | --- | --- | --- | --- | --- | --- | --- | --- | --- | --- | --- | --- | --- | --- |
| **Gene** | **Variant** | **Genotype** | **Controls**  **N** | **Cases**  **N** | **Cases**  **%** | **P^a^** | **No sleep**  **Meds N** | **Sleep**  **Meds N** | **Sleep**  **Meds %** | **P^a^** | **Controls**  **N** | **Cases**  **N** | **Cases**  **%** | **P^a^** |
| *ADRB1* | A187V | C/C | 164,526 | 5,921 | 3.47 | 0.310 | 155,977 | 14,467 | 8.49 | 0.386 | 170,420 | 27 | 0.02 | 1.000 |
|  |  | C/T | 66 | 4 | 5.71 |  | 62 | 8 | 11.43 |  | 70 | 0 | 0.00 |  |
| *BHLHE41 (DEC2)* | P384R | G/G | 164,577 | 5,921 | 3.47 | 0.298 | 156,023 | 14,472 | 8.49 | 0.588 | 170,471 | 27 | 0.02 | 1.000 |
|  |  | G/C | 9 | 1 | 10.00 |  | 9 | 1 | 10.00 |  | 10 | 0 | 0.00 |  |
| *GRM1* | S458A | T/T | 164,526 | 5,923 | 3.47 | 1.000 | 155,976 | 14,470 | 8.49 | 1.000 | 170,422 | 27 | 0.02 | 1.000 |
|  |  | T/G | 65 | 2 | 2.99 |  | 62 | 5 | 7.46 |  | 67 | 0 | 0.00 |  |
|  | A889T | A/A | 164,586 | 5,925 | 3.47 | 1.000 | 156,033 | 14,475 | 8.49 | 1.000 | 170,484 | 27 | 0.02 | 1.000 |
|  |  | A/T | 3 | 0 | 0.00 |  | 3 | 0 | 0.00 |  | 3 | 0 | 0.00 |  |
| *PER3* | P415A | C/C | 163,079 | 5,862 | 3.47 | 0.240 | 154,589 | 14,348 | 8.49 | 0.413 | 168914 | 27 | 0.02 | 1.000 |
|  |  | C/G | 1,505 | 62 | 3.96 |  | 1,442 | 123 | 7.86 |  | 1567 | 0 | 0.00 |  |
|  |  | G/G | 6 | 1 | 14.29 |  | 6 | 1 | 14.29 |  | 7 | 0 | 0.00 |  |
|  | H417R | A/A | 163,078 | 5,862 | 3.47 | 0.268 | 154,589 | 14,348 | 8.49 | 0.525 | 168913 | 27 | 0.02 | 1.000 |
|  |  | A/G | 1,507 | 62 | 3.95 |  | 1,444 | 125 | 7.97 |  | 1569 | 0 | 0.00 |  |
|  |  | G/G | 6 | 1 | 14.29 |  | 6 | 1 | 14.29 |  | 7 | 0 | 0.00 |  |
| *CRY2* | A260T | G/G | 164,555 | 5,924 | 3.47 | 1.000 | 156,008 | 14,468 | 8.49 | 0.044 | 170452 | 27 | 0.02 | 1.000 |
|  |  | G/A | 38 | 1 | 5.71 |  | 32 | 7 | 17.95 |  | 39 | 0 | 0.00 |  |
| *TIMELESS* | R1081X | G/G | 164,580 | 5,925 | 3.47 | 1.000 | 156,027 | 14,475 | 8.49 | 1.000 | 170478 | 27 | 0.02 | 1.000 |
|  |  | G/A | 6 | 0 | 5.71 |  | 6 | 0 | 0.00 |  | 6 | 0 | 0.00 |  |
| *CRY1* | c.1657+3A>C | T/T | 163,148 | 5,879 | 3.47 | 0.477 | 154,669 | 14,355 | 8.49 | 0.575 | 169000 | 27 | 0.02 | 1.000 |
|  |  | T/G | 1,435 | 46 | 3.47 |  | 1,362 | 119 | 8.04 |  | 1481 | 0 | 0.00 |  |
|  |  | G/G | 9 | 0 | 5.71 |  | 8 | 1 | 11.11 |  | 9 | 0 | 0.00 |  |
| *CSNK1D* | T44A | T/T | 164,592 | 5,925 | 3.47 | 1.000 | 156,039 | 14,475 | 8.49 | 1.000 | 170490 | 27 | 0.02 | 1.000 |
|  |  | T/C | 1 | 0 | 5.71 |  | 1 | 0 | 0.00 |  | 1 | 0 | 0.00 |  |

^a^P-value derived from 2-sided Fisher’s exact test. Homozygous carriers for minor alleles were combined with heterozygous carriers prior to performing Fisher’s exact test.

Supplementary Table 2. Summary statistics of self-reported sleep duration in the UK Biobank (Field 1160) for carriers of variants previously described as causal for familial natural short sleep.

| **Gene** | **Variant** | **REF/ALT^a^** | **Study** | **Genotype** | **N** | **Minimum** | **Maximum** | **Mean** | **SD^b^** | **P^c^** |
| --- | --- | --- | --- | --- | --- | --- | --- | --- | --- | --- |
| *ADRB1* | A187V | C/T | UKB | C/C | 166,291 | 1 | 12 | 7.17 | 1.07 | 0.615 |
|  |  |  |  | C/T | 69 | 5 | 12 | 7.10 | 1.03 |  |
| *BHLHE41 / DEC2* | P384R | G/C | UKB | G/G | 166,283 | 1 | 12 | 7.17 | 1.07 | 0.694 |
|  |  |  |  | G/C | 10 | 5 | 9 | 7.30 | 1.06 |  |
| *GRM1* | S458A | T/G | UKB | T/T | 166,290 | 1 | 12 | 7.17 | 1.07 | 0.807 |
|  |  |  |  | T/G | 67 | 4 | 10 | 7.13 | 1.15 |  |
|  |  |  | Finnish | T/T | 12027 | 3 | 15 | 7.4 | 1.14 | 0.390 |
|  |  |  |  | T/G | 6 | 5 | 9 | 7 | 1.41 |  |
|  | A889T | A/T | UKB | A/A | 166,288 | 1 | 12 | 7.17 | 1.07 | 0.179 |
|  |  |  |  | A/T | 3 | 8 | 8 | 8.00 | 0.00 |  |

^a^Reference and alternate allele relative to reference genome; ^b^Standard Deviation; ^c^P-value derived from 2-sided t-test.

Supplementary Table 3. Summary statistics of dichotomised self-reported sleep data in the UK Biobank for carriers of variants previously described as causal for familial natural short sleep. Data unavailable for self-reported sleep of ≤5 hours, ≤4 hours and 4-6 hours in the Finnish study.

|  |  |  |  |  | **Short Sleep ( ≤6 hours)** | | | | **Short Sleep (≤5 hours)** | | | | **Short Sleep (≤4 hours)** | | | | **Short Sleep (4 to 6 hours)** | | | |
| --- | --- | --- | --- | --- | --- | --- | --- | --- | --- | --- | --- | --- | --- | --- | --- | --- | --- | --- | --- | --- |
| **Gene** | **Variant** | **REF/**  **ALT^a^** | **Study** | **Genotype** | **%**  **cases** | **N**  **cases** | **N**  **controls** | **P^b^** | **%**  **cases** | **N**  **cases** | **N**  **controls** | **P^b^** | **%**  **cases** | **N**  **cases** | **N**  **controls** | **P^b^** | **%**  **cases** | **N**  **cases** | **N**  **controls** | **P^b^** |
| *ADRB1* | A187V | C/T | UKB | C/C | 23.74 | 39,484 | 126,807 | 1.000 | 5.08 | 8,446 | 157,845 | 1.000 | 1.01 | 1,679 | 164,612 | 1.000 | 23.56 | 39,176 | 127,115 | 1.000 |
|  |  |  |  | C/T | 23.19 | 16 | 53 |  | 4.35 | 3 | 66 |  | 0.00 | 0 | 69 |  | 23.19 | 16 | 53 |  |
| *BHLHE41*  */ DEC2* | P384R | G/C | UKB | G/G | 23.74 | 39,483 | 126,800 | 0.469 | 5.08 | 8,446 | 157,837 | 0.406 | 1.01 | 1,679 | 164,604 | 1.000 | 23.56 | 39,175 | 127,108 | 0.470 |
|  |  |  |  | G/C | 10.00 | 1 | 9 |  | 10.00 | 1 | 9 |  | 0.00 | 0 | 10 |  | 10.00 | 1 | 9 |  |
| *GRM1* | S458A | T/G | UKB | T/T | 23.74 | 39,484 | 126,806 | 0.389 | 5.08 | 8,446 | 157,844 | 0.393 | 1.01 | 1,679 | 164,611 | 0.494 | 23.56 | 39,176 | 127,114 | 0.387 |
|  |  |  |  | T/G | 28.36 | 19 | 48 |  | 7.46 | 5 | 62 |  | 1.49 | 1 | 66 |  | 28.36 | 19 | 48 |  |
|  |  |  | Finnish | T/T | 17.88 | 2,151 | 9,876 | 0.290 | NA | NA | NA | NA | NA | NA | NA | NA | NA | NA | NA | NA |
|  |  |  |  | T/G | 33.33 | 2 | 4 |  | NA | NA | NA |  | NA | NA | NA |  | NA | NA | NA |  |
|  | A889T | A/T | UKB | A/A | 23.75 | 39,485 | 126,803 | 1.000 | 5.08 | 8,446 | 157,842 | 1.000 | 1.01 | 1,679 | 164,609 | 1.000 | 23.56 | 39,177 | 127,111 | 1.000 |
|  |  |  |  | A/T | 0.00 | 0 | 3 |  | 0.00 | 0 | 3 |  | 0.00 | 0 | 3 |  | 0.00 | 0 | 3 |  |

^a^Reference and alternate allele relative to reference genome; ^b^P-value derived from 2-sided Fisher’s exact-test.

Supplementary Table 4. Summary statistics of accelerometer-derived estimates of sleep duration in UK Biobank for carriers of variants previously described as casual for familial natural short sleep. No carriers of the *GRM1* A889T variant remained among individuals from UK Biobank who had worn an accelerometer.

|  |  |  |  | **All Nights (hours)** | | | | | **Weeknights (hours)** | | | | | **Weekend Nights (hours)** | | | | |
| --- | --- | --- | --- | --- | --- | --- | --- | --- | --- | --- | --- | --- | --- | --- | --- | --- | --- | --- |
| **Gene** | **Variant** | **REF/**  **ALT^a^** | **Genotype** | **N** | **Min^b^** | **Max^c^** | **Mean (SD^d^)** | **P^e^** | **N** | **Min^b^** | **Max^c^** | **Mean (SD^d^)** | **P^e^** | **N** | **Min^b^** | **Max^c^** | **Mean (SD^d^)** | **P^e^** |
| *ADRB1* | A187V | C/T | C/C | 34,168 | 1.63 | 11.87 | 7.30 (0.86) | 0.197 | 34,134 | 1.35 | 11.87 | 7.25 (0.92) | 0.178 | 33,271 | 1.01 | 11.63 | 7.44 (1.09) | 0.500 |
|  |  |  | C/T | 15 | 6.89 | 8.83 | 7.59 (0.49) |  | 15 | 6.90 | 8.73 | 7.57 (0.47) |  | 15 | 6.50 | 9.03 | 7.63 (0.81) |  |
| *BHLHE41 / DEC2* | P384R | G/C | G/G | 34,167 | 1.63 | 11.87 | 7.30 (0.86) | 0.624 | 34,133 | 1.35 | 11.87 | 7.25 (0.92) | 0.484 | 33,270 | 1.01 | 11.63 | 7.44 (1.09) | 0.858 |
|  |  |  | G/C | 4 | 6.26 | 8.25 | 7.51 (0.92) |  | 4 | 6.25 | 8.29 | 7.57 (0.92) |  | 4 | 6.27 | 8.60 | 7.35 (1.06) |  |
| *GRM1* | S458A | T/G | T/T | 34,166 | 1.63 | 11.87 | 7.30 (0.86) | 0.915 | 34,132 | 1.35 | 11.87 | 7.25 (0.92) | 0.590 | 33,269 | 1.01 | 11.63 | 7.44 (1.09) | 0.331 |
|  |  |  | T/G | 10 | 6.48 | 8.30 | 7.33 (0.76) |  | 10 | 6.55 | 8.34 | 7.40 (0.79) |  | 10 | 5.07 | 8.87 | 7.11 (1.14) |  |
| *GRM1* | A889T | A/T | A/A | 34,167 | 1.63 | 11.87 | 7.30 (0.86) | NA | 34,133 | 1.35 | 11.87 | 7.25 (0.92) | NA | 33,270 | 1.01 | 11.63 | 7.44 (1.09) | NA |
|  |  |  | A/T | 0 | NA | NA | NA |  | 0 | NA | NA | NA |  | 0 | NA | NA | NA |  |

^a^Reference and alternate allele relative to reference genome; ^b^Minimum; ^c^Maximum; ^d^Standard deviation; ^e^P-value from 2-sided t-test.

Supplementary Table 5. Summary statistics of “Morningness” across genotype groups for variants previously reported as causal for familial advanced sleep phase. Data on being “more or definitely a morning person” unavailable in the Finnish studies.

|  |  |  |  | **Definitely a morning person** | | | | **More or definitely a morning person** | | | |
| --- | --- | --- | --- | --- | --- | --- | --- | --- | --- | --- | --- |
| **Gene** | **Variant** | **Study** | **Genotype** | **%**  **cases** | **N**  **cases** | **N**  **controls** | **P^a^** | **%**  **cases** | **N**  **cases** | **N**  **controls** | **P^a^** |
| *PER3* | P415A | UKB | C/C | 23.77 | 39,655 | 127,177 | <0.0001 | 56.24 | 93,827 | 73,005 | <0.0001 |
|  |  |  | C/G | 29.58 | 463 | 1,102 |  | 62.94 | 985 | 580 |  |
|  |  |  | G/G | 42.86 | 3 | 4 |  | 71.43 | 5 | 2 |  |
|  |  | Finnish | C/C | 22.6 | 1,795 | 6,146 | 0.293 | NA | NA | NA | NA |
|  |  |  | C/G | 24.9 | 109 | 328 |  | NA | NA | NA |  |
|  |  |  | G/G | 0 | 0 | 3 |  | NA | NA | NA |  |
|  | H417R | UKB | A/A | 23.77 | 39,655 | 127,177 | <0.0001 | 56.24 | 93,827 | 73,005 | <0.0001 |
|  |  |  | A/G | 29.55 | 463 | 1,104 |  | 62.86 | 985 | 582 |  |
|  |  |  | G/G | 42.86 | 3 | 4 |  | 71.43 | 5 | 2 |  |
|  |  | Finnish | A/A | 22.6 | 1,797 | 6,148 | 0.268 | NA | NA | NA | NA |
|  |  |  | A/G | 25 | 109 | 327 |  | NA | NA | NA |  |
|  |  |  | G/G | 16.7 | 1 | 5 |  | NA | NA | NA |  |
| *CRY2* | A260T | UKB | G/G | 23.77 | 39,655 | 127,179 | 0.340 | 56.24 | 93,827 | 73,007 | 0.327 |
|  |  |  | G/A | 15.79 | 6 | 32 |  | 47.37 | 18 | 20 |  |
| *TIMELESS* | R1081X | UKB | G/G | 23.77 | 39,652 | 127,175 | 1.000 | 56.24 | 93,822 | 73,005 | 0.175 |
|  |  |  | G/A | 20.00 | 1 | 4 |  | 20 | 1 | 4 |  |

^a^P-value derived from 2-sided Fisher’s exact test. Homozygous carriers for minor alleles were combined with heterozygous carriers prior to performing Fisher’s exact test.

### Supplementary Table 6. Summary statistics of sleep-midpoint estimated from accelerometer data in UK Biobank and MESA across genotype groups for variants previously reported as causal for familial advanced sleep phase.

|  |  |  |  | **Average for All Nights** | | | | | **Average for Weeknights** | | | | | **Average for Weekend Nights** | | | | |
| --- | --- | --- | --- | --- | --- | --- | --- | --- | --- | --- | --- | --- | --- | --- | --- | --- | --- | --- |
| **Gene** | **Variant** | **Study** | **Genotype** | **N** | **Min^a^** | **Max^b^** | **Mean (SD^c^)** | **P^d^** | **N** | **Min^a^** | **Max^b^** | **Mean (SD^c^)** | **P^d^** | **N** | **Min^a^** | **Max^b^** | **Mean (SD^c^)** | **P^d^** |
| *PER3* | P415A | UKB | C/C | 33,908 | 23.37 | 30.59 | 27.01 (0.85) | 0.014 | 33,874 | 23.01 | 30.59 | 26.92 (0.91) | 0.005 | 33,022 | 21.69 | 32.68 | 27.27 (1.25) | 0.256 |
|  |  |  | C/G | 338 | 19.45 | 29.64 | 26.90 (0.96) |  | 337 | 19.22 | 29.42 | 26.77 (1.03) |  | 333 | 18.92 | 30.99 | 27.19 (1.48) |  |
|  |  |  | G/G | 1 | 27.60 | 27.60 | 27.60 |  | 1 | 27.39 | 27.39 | 27.39 |  | 1 | 28.12 | 28.12 | 28.12 |  |
|  |  | MESA | C/C | 1,925 | 13.15 | 34.92 | 27.05 (2.16) | 0.900 | 1,924 | 13.00 | 34.69 | 27.02 (2.02) | 0.888 | 1,915 | 13.00 | 33.92 | 27.46 (1.7) | 0.123 |
|  |  |  | C/G | 10 | 26.12 | 27.98 | 26.96 (0.67) |  | 10 | 26.08 | 28.64 | 27.11 (0.84) |  | 10 | 25.46 | 27.80 | 26.63 (0.66) |  |
|  | H417R | UKB | A/A | 33,907 | 23.37 | 30.59 | 27.01 (0.85) | 0.018 | 33,873 | 23.01 | 30.59 | 26.92 (0.91) | 0.006 | 33,021 | 21.69 | 32.68 | 27.27 (1.25) | 0.272 |
|  |  |  | A/G | 339 | 19.45 | 29.64 | 26.90 (0.96) |  | 338 | 19.22 | 29.42 | 26.78 (1.03) |  | 334 | 18.92 | 30.99 | 27.19 (1.48) |  |
|  |  |  | G/G | 1 | 27.60 | 27.60 | 27.60 |  | 1 | 27.39 | 27.39 | 27.39 |  | 1 | 28.12 | 28.12 | 28.12 |  |
|  |  | MESA | A/A | 1,925 | 13.15 | 34.92 | 27.05 (2.16) | 0.900 | 1,924 | 13.00 | 34.69 | 27.02 (2.02) | 0.888 | 1,915 | 13.00 | 33.92 | 27.46 (1.7) | 0.123 |
|  |  |  | A/G | 10 | 26.12 | 27.98 | 26.96 (0.67) |  | 10 | 26.08 | 28.64 | 27.11 (0.84) |  | 10 | 25.46 | 27.80 | 26.63 (0.66) |  |
| *CRY2* | A260T | UKB | G/G | 33,908 | 23.37 | 30.59 | 27.01 (0.85) | 0.040 | 33,874 | 23.01 | 30.59 | 26.92 (0.91) | 0.142 | 33,022 | 21.69 | 32.68 | 27.27 (1.25) | 0.005 |
|  |  |  | G/A | 4 | 26.63 | 29.34 | 27.88 (1.14) |  | 4 | 26.63 | 28.72 | 27.59 (0.91) |  | 3 | 28.38 | 30.89 | 29.29 (1.39) |  |
| *TIMELESS* | R1081X | UKB | G/G | 33,907 | 23.37 | 30.59 | 27.01 (0.85) | NA | 33,873 | 23.01 | 30.59 | 26.92 (0.91) | NA | 33,021 | 21.69 | 32.68 | 27.27 (1.25) | NA |
|  |  |  | G/A | 0 | NA | NA | NA |  | 0 | NA | NA | NA |  | 0 | NA | NA | NA |  |

^a^Minimum; ^b^Maximum; ^c^Standard Deviation; ^d^P-value from 2-sided t-test comparing means and standard deviations of homozygous carriers for minor alleles with heterozygotes.

### Supplementary Table 7. Summary statistics of L5-midpoint timing estimated from accelerometer data in UK Biobank across genotype groups for variants previously reported as causal for familial advanced sleep phase.

|  |  |  | **Average for All Nights** | | | | | **Average for Weeknights** | | | | | **Average for Weekend Nights** | | | | |
| --- | --- | --- | --- | --- | --- | --- | --- | --- | --- | --- | --- | --- | --- | --- | --- | --- | --- |
| **Gene** | **Variant** | **Genotype** | **N** | **Min^a^** | **Max^b^** | **Mean (SD^c^)** | **P^d^** | **N** | **Min^a^** | **Max^b^** | **Mean (SD^c^)** | **P^d^** | **N** | **Min^a^** | **Max^b^** | **Mean (SD^c^)** | **P^d^** |
| *PER3* | P415A | C/C | 33,998 | 23.08 | 31.51 | 27.32 (0.99) | 0.053 | 33,964 | 22.79 | 31.80 | 27.29 (1.05) | 0.053 | 32,636 | 21.50 | 33.32 | 27.42 (1.36) | 0.076 |
|  |  | C/G | 338 | 21.34 | 31.43 | 27.21 (1.05) |  | 337 | 20.09 | 31.53 | 27.18 (1.16) |  | 325 | 13.00 | 30.92 | 27.28 (1.57) |  |
|  |  | G/G | 1 | 28.01 | 28.01 | 28.01 |  | 1 | 27.30 | 27.30 | 27.30 |  | 1 | 29.44 | 29.44 | 29.44 |  |
|  | H417R | A/A | 33,997 | 23.08 | 31.51 | 27.32 (0.99) | 0.060 | 33,963 | 22.79 | 31.80 | 27.29 (1.05) | 0.060 | 32,635 | 21.50 | 33.32 | 27.42 (1.36) | 0.076 |
|  |  | A/G | 339 | 21.34 | 31.43 | 27.21 (1.05) |  | 338 | 20.09 | 31.53 | 27.19 (1.16) |  | 325 | 13.00 | 30.92 | 27.28 (1.57) |  |
|  |  | G/G | 1 | 28.01 | 28.01 | 28.01 |  | 1 | 27.30 | 27.30 | 27.30 |  | 1 | 29.44 | 29.44 | 29.44 |  |
| *CRY2* | A260T | G/G | 33,998 | 23.08 | 31.51 | 27.32 (0.99) | 0.092 | 33,964 | 22.79 | 31.80 | 27.29 (1.05) | 0.311 | 32,636 | 21.50 | 33.32 | 27.42 (1.36) | 0.031 |
|  |  | G/A | 4 | 27.32 | 28.67 | 28.15 (0.58) |  | 4 | 26.91 | 28.67 | 27.82 (0.73) |  | 3 | 28.98 | 29.31 | 29.12 (0.17) |  |
| *TIMELESS* | R1081X | G/G | 33,997 | 23.08 | 31.51 | 27.32 (0.99) | NA | 33,963 | 22.79 | 31.80 | 27.29 (1.05) | NA | 32,635 | 21.50 | 33.32 | 27.42 (1.36) | NA |
|  |  | G/A | 0 | NA | NA | NA |  | 0 | NA | NA | NA |  | 0 | NA | NA | NA |  |

^a^Minimum; ^b^Maximum; ^c^Standard Deviation; ^d^P-value from 2-sided t-test comparing means and standard deviations of homozygous carriers for minor alleles with heterozygotes.

Supplementary Table 8. Summary statistics of “eveningness” across genotype groups for variants previously reported as causal for delayed sleep phase. Data on being “more or definitely an evening person” unavailable in the Finnish studies.

|  |  |  |  | **Definitely an evening person** | | | | **More or definitely an evening person** | | | |
| --- | --- | --- | --- | --- | --- | --- | --- | --- | --- | --- | --- |
| **Gene** | **Variant** | **Study** | **Genotype** | **%**  **cases** | **N**  **cases** | **N**  **controls** | **P^a^** | **%**  **cases** | **N**  **cases** | **N**  **controls** | **P^a^** |
| *CRY1* | c.1657+3A>C | UKB | T/T | 7.92 | 13,212 | 153,621 | 0.003 | 33.48 | 55,862 | 110,971 | 0.001 |
|  |  |  | T/G | 10.07 | 149 | 1,331 |  | 37.91 | 561 | 919 |  |
|  |  |  | G/G | 11.11 | 1 | 8 |  | 22.22 | 2 | 7 |  |
|  |  | Finnish | T/T | 10 | 284 | 2,554 | 1.000 | NA | NA | NA | NA |
|  |  |  | T/G | 0 | 0 | 4 |  | NA | NA | NA |  |

^a^P-value from 2-sided Fisher’s exact test. Homozygous carriers for minor alleles were combined with heterozygous carriers prior to performing Fisher’s exact test.

### Supplementary Table 9. Summary statistics of sleep-midpoint estimated from accelerometer data in UK Biobank and MESA across genotype groups for variants previously reported as causal for delayed sleep phase.

|  |  |  |  | **Average for All Nights** | | | | | **Average for Weeknights** | | | | | **Average for Weekend Nights** | | | | |
| --- | --- | --- | --- | --- | --- | --- | --- | --- | --- | --- | --- | --- | --- | --- | --- | --- | --- | --- |
| **Gene** | **Variant** | **Study** | **Genotype** | **N** | **Min^a^** | **Max^b^** | **Mean (SD^c^)** | **P^d^** | **N** | **Min^a^** | **Max^b^** | **Mean (SD^c^)** | **P^d^** | **N** | **Min^a^** | **Max^b^** | **Mean (SD^c^)** | **P^d^** |
| *CRY1* | c.1657+3A>C | UKB | T/T | 33,908 | 23.37 | 30.59 | 27.01 (0.85) | 0.059 | 33,874 | 23.01 | 30.59 | 26.92 (0.91) | 0.038 | 33,022 | 21.69 | 32.68 | 27.27 (1.25) | 0.325 |
|  |  |  | T/G | 318 | 24.28 | 29.60 | 27.11 (0.88) |  | 318 | 23.51 | 29.44 | 27.03 (0.94) |  | 301 | 18.95 | 30.86 | 27.35 (1.46) |  |
|  |  |  | G/G | 4 | 25.49 | 27.38 | 26.5 (0.9) |  | 4 | 25.81 | 27.54 | 26.64 (0.73) |  | 4 | 23.25 | 27.77 | 26.25 (2.03) |  |
|  |  | MESA | T/T | 1914 | 13.19 | 34.92 | 27.05 (2.14) | 0.568 | 1,913 | 13.00 | 34.69 | 27.03 (2.01) | 0.668 | 1,904 | 13.00 | 33.92 | 27.45 (1.7) | 0.668 |
|  |  |  | T/G | 21 | 13.15 | 30.10 | 26.78 (3.32) |  | 21 | 15.59 | 29.85 | 26.84 (2.78) |  | 21 | 25.02 | 30.80 | 27.61 (1.54) |  |

^a^Minimum; ^b^Maximum; ^c^Standard Deviation; ^d^P-value from 2-sided t-test comparing means and standard deviations of homozygous carriers for minor alleles with heterozygotes.

### Supplementary Table 10. Summary statistics of L5-midpoint timing estimated from accelerometer data in UK Biobank a across genotype groups for variants previously reported as causal for delayed sleep phase.

|  |  |  | **Average for All Nights** | | | | | **Average for Weeknights** | | | | | **Average for Weekend Nights** | | | | |
| --- | --- | --- | --- | --- | --- | --- | --- | --- | --- | --- | --- | --- | --- | --- | --- | --- | --- |
| **Gene** | **Variant** | **Genotype** | **N** | **Min^a^** | **Max^b^** | **Mean (SD^c^)** | **P^d^** | **N** | **Min^a^** | **Max^b^** | **Mean (SD^c^)** | **P^d^** | **N** | **Min^a^** | **Max^b^** | **Mean (SD^c^)** | **P^d^** |
| *CRY1* | c.1657+3A>C | T/T | 33,998 | 23.08 | 31.51 | 27.32 (0.99) | 0.132 | 33,964 | 22.79 | 31.80 | 27.29 (1.05) | 0.163 | 32,636 | 21.50 | 33.32 | 27.42 (1.36) | 0.335 |
|  |  | T/G | 318 | 24.06 | 30.38 | 27.41 (1.00) |  | 318 | 23.54 | 30.11 | 27.38 (1.06) |  | 305 | 17.84 | 31.67 | 27.51 (1.39) |  |
|  |  | G/G | 4 | 25.08 | 28.44 | 26.5 (1.48) |  | 4 | 25.67 | 28.63 | 26.72 (1.33) |  | 3 | 22.82 | 28.08 | 26.05 (2.82) |  |

^a^Minimum; ^b^Maximum; ^c^Standard Deviation; ^d^P-value from 2-sided t-test comparing means and standard deviations of homozygous carriers for minor alleles with heterozygotes.

### Supplementary Table 11. P-values from burden testing of rare (MAF < 0.01%) loss-of-function and missense variants in genes outlined in this paper on self-reported sleep duration in UK Biobank.

| **Gene** | **Canonical**  **Transcript** | **Reported**  **Trait** | **Variant class** | **Sleep**  **Duration^e^** | **Sleep**  **Duration^f^** | **Sleep Duration**  **4 to 6 hours** | **Sleep Duration**  **1 to 6 hours** | **Sleep Duration**  **1 to 5 hours** | **Sleep Duration**  **1 to 4 hours** |
| --- | --- | --- | --- | --- | --- | --- | --- | --- | --- |
| *GRM1* | ENST00000361719 | FNSS^a^ | LoF^d^ | 0.872 | 0.690 | 0.556 | 0.583 | 0.183 | 0.629 |
|  |  |  | Missense | 0.413 | 0.401 | 0.907 | 0.978 | 0.964 | 0.017 |
| *NPSR1* | ENST00000359791 | FNSS^a^ | LoF^d^ | 0.431 | 0.863 | 0.647 | 0.471 | 0.022 | 0.208 |
|  |  |  | Missense | 0.445 | 0.790 | 0.560 | 0.479 | 0.493 | 0.832 |
| *ADRB1* | ENST00000369295 | FNSS^a^ | LoF^d^ | 0.204 | 0.266 | 0.340 | 0.336 | 0.458 | 0.789 |
|  |  |  | Missense | 0.150 | 0.238 | 0.076 | 0.091 | 0.390 | 0.934 |
| *BHLHE41* | ENST00000242728 | FNSS^a^ | LoF^d^ | 0.676 | 0.484 | 0.291 | 0.301 | 0.924 | 0.051 |
|  |  |  | Missense | 0.530 | 0.475 | 0.034 | 0.033 | 0.905 | 0.520 |
| *CRY1* | ENST00000008527 | DSPD^b^ | LoF^d^ | 0.713 | 0.898 | 0.636 | 0.580 | 0.653 | 0.353 |
|  |  |  | Missense | 0.217 | 0.292 | 0.086 | 0.072 | 0.199 | 0.583 |
| *PER3* | ENST00000361923 | FASP^c^ | LoF^d^ | 0.831 | 0.542 | 0.473 | 0.490 | 0.568 | 0.822 |
|  |  |  | Missense | 0.035 | 0.016 | 0.109 | 0.166 | 0.708 | 0.939 |
| *PER2* | ENST00000254657 | FASP^c^ | LoF^d^ | 0.522 | 0.400 | 0.909 | 0.924 | 0.101 | 0.412 |
|  |  |  | Missense | 0.819 | 0.796 | 0.610 | 0.532 | 0.299 | 0.930 |
| *CRY2* | ENST00000443527 | FASP^c^ | LoF^d^ | 0.417 | 0.611 | 0.197 | 0.192 | 0.830 | 0.661 |
|  |  |  | Missense | 0.053 | 0.085 | 0.141 | 0.161 | 0.389 | 0.687 |
| *TIMELESS* | ENST00000553532 | FASP^c^ | LoF^d^ | 0.659 | 0.871 | 0.296 | 0.336 | 0.261 | 0.715 |
|  |  |  | Missense | 0.420 | 0.365 | 0.571 | 0.439 | 0.559 | 0.789 |
| *CSNK1D* | ENST00000314028 | FASP^c^ | LoF^d^ | 0.124 | 0.042 | 0.004 | 0.004 | 0.479 | 0.807 |
|  |  |  | Missense | 0.701 | 0.659 | 0.653 | 0.624 | 0.198 | 0.053 |

^a^FNSS=familial natural short sleep; ^b^DSP=delayed sleep phase disorder; ^c^FASP=familial advanced sleeo phase; ^d^LoF=loss-of-function; ^e^Sleep duration analysed on original unit scale; ^f^Sleep duration inverse-normalised prior to analysis.

### Supplementary Table 12. P-values from burden testing of rare (MAF < 0.01%) loss-of-function and missense variants in genes outlined in this paper on chronotype in UK Biobank.

| **Gene** | **Canonical**  **Transcript** | **Reported**  **Trait** | **Variant Class** | **Chronotype** | | **Definitely an**  **Evening Person** | | **Definitely a**  **Morning Person** | | **More or Definitely**  **an Evening Person** | | **More or Definitely**  **a Morning Person** |
| --- | --- | --- | --- | --- | --- | --- | --- | --- | --- | --- | --- | --- |
| *GRM1* | ENST00000361719 | FNSS^a^ | LoF^d^ | 0.685 | 0.508 | | 0.711 | | 0.689 | | 0.402 | |
|  |  |  | Missense | 0.676 | 0.684 | | 0.439 | | 0.733 | | 0.592 | |
| *NPSR1* | ENST00000359791 | FNSS^a^ | LoF^d^ | 0.911 | 0.363 | | 0.693 | | 0.770 | | 0.440 | |
|  |  |  | Missense | 0.686 | 0.517 | | 0.795 | | 0.884 | | 0.385 | |
| *ADRB1* | ENST00000369295 | FNSS^a^ | LoF^d^ | 0.055 | 0.349 | | 0.076 | | 0.016 | | 0.066 | |
|  |  |  | Missense | 0.682 | 0.415 | | 0.422 | | 0.419 | | 0.725 | |
| *BHLHE41* | ENST00000242728 | FNSS^a^ | LoF^d^ | 0.956 | 0.261 | | 0.172 | | 0.654 | | 0.981 | |
|  |  |  | Missense | 0.585 | 0.593 | | 0.933 | | 0.693 | | 0.145 | |
| *CRY1* | ENST00000008527 | DSPD^b^ | LoF^d^ | 0.326 | 0.326 | | 0.214 | | 0.541 | | 0.534 | |
|  |  |  | Missense | 0.405 | 0.096 | | 0.965 | | 0.234 | | 0.767 | |
| *PER3* | ENST00000361923 | FASP^c^ | LoF^d^ | 0.009 | 0.791 | | 0.005 | | 0.018 | | 0.007 | |
|  |  |  | Missense | 0.681 | 0.091 | | 0.021 | | 0.183 | | 0.353 | |
| *PER2* | ENST00000254657 | FASP^c^ | LoF^d^ | 1.3E-09 | 0.004 | | 3.8E-08 | | 3.4E-06 | | 5.0E-09 | |
|  |  |  | Missense | 0.345 | 0.277 | | 0.341 | | 0.811 | | 0.471 | |
| *CRY2* | ENST00000443527 | FASP^c^ | LoF^d^ | 0.015 | 0.856 | | 0.008 | | 0.085 | | 0.006 | |
|  |  |  | Missense | 0.816 | 0.492 | | 0.318 | | 0.808 | | 0.991 | |
| *TIMELESS* | ENST00000553532 | FASP^c^ | LoF^d^ | 0.512 | 0.471 | | 0.761 | | 0.926 | | 0.096 | |
|  |  |  | Missense | 0.834 | 0.250 | | 0.245 | | 0.713 | | 0.775 | |
| *CSNK1D* | ENST00000314028 | FASP^c^ | LoF^d^ | 0.199 | 0.966 | | 0.311 | | 0.154 | | 0.232 | |
|  |  |  | Missense | 0.296 | 0.150 | | 0.358 | | 0.496 | | 0.490 | |

^a^FNSS=familial natural short sleep; ^b^DSP=delayed sleep phase disorder; ^c^FASP=familial advanced sleeo phase; ^d^LoF=loss-of-function.

Supplementary Table 13. P-values from burden testing of rare (MAF < 0.01%) loss-of-function and missense variants in genes previously reported to harbour variants causal for disruptive sleep duration or timing on accelerometer estimates of sleep duration in UK Biobank. There were no remaining loss-of-function carriers for GRM1, ADRB1 and CRY2 within the subset of individuals from UK Biobank who wore an accelerometer.

| **Gene** | **Canonical**  **Transcript** | **Reported**  **Trait** | **Variant Class** | **Sleep**  **Duration^e^** | **Sleep**  **Duration^f^** | **Sleep Duration**  **4 to 6 hours** | **Sleep Duration**  **1 to 6 hours** | **Sleep Duration**  **1 to 5 hours** | **Sleep Duration**  **1 to 4 hours** |
| --- | --- | --- | --- | --- | --- | --- | --- | --- | --- |
| *GRM1* | ENST00000361719 | FNSS^a^ | LoF^d^ | NA | NA | NA | NA | NA | NA |
|  |  |  | Missense | 0.862 | 0.750 | 0.196 | 0.159 | 0.409 | 0.470 |
| *NPSR1* | ENST00000359791 | FNSS^a^ | LoF^d^ | 0.044 | 0.030 | 0.055 | 0.108 | 0.768 | 0.896 |
|  |  |  | Missense | 0.950 | 0.767 | 0.710 | 0.770 | 0.266 | 0.522 |
| *ADRB1* | ENST00000369295 | FNSS^a^ | LoF^d^ | NA | NA | NA | NA | NA | NA |
|  |  |  | Missense | 0.729 | 0.849 | 0.053 | 0.041 | 0.439 | 0.565 |
| *BHLHE41* | ENST00000242728 | FNSS^a^ | LoF^d^ | 0.052 | 0.053 | 0.482 | 0.478 | 0.780 | 0.926 |
|  |  |  | Missense | 0.865 | 0.722 | 0.330 | 0.311 | 0.171 | 0.559 |
| *CRY1* | ENST00000008527 | DSPD^b^ | LoF^d^ | 0.880 | 0.925 | 0.263 | 0.254 | 0.740 | 0.884 |
|  |  |  | Missense | 0.914 | 0.920 | 0.973 | 0.912 | 0.681 | 0.660 |
| *PER3* | ENST00000361923 | FASP^c^ | LoF^d^ | 0.622 | 0.590 | 0.943 | 0.883 | 0.447 | 0.625 |
|  |  |  | Missense | 0.481 | 0.517 | 0.220 | 0.192 | 0.222 | 0.235 |
| *PER2* | ENST00000254657 | FASP^c^ | LoF^d^ | 0.178 | 0.192 | 0.321 | 0.318 | 0.037 | 0.847 |
|  |  |  | Missense | 0.878 | 0.724 | 0.753 | 0.977 | 0.332 | 0.337 |
| *CRY2* | ENST00000443527 | FASP^c^ | LoF^d^ | NA | NA | NA | NA | NA | NA |
|  |  |  | Missense | 0.575 | 0.687 | 0.954 | 0.811 | 0.506 | 0.514 |
| *TIMELESS* | ENST00000553532 | FASP^c^ | LoF^d^ | 0.001 | 8.4E-04 | 0.438 | 0.341 | 0.376 | 0.752 |
|  |  |  | Missense | 0.974 | 0.779 | 0.355 | 0.198 | 0.266 | 0.512 |
| *CSNK1D* | ENST00000314028 | FASP^c^ | LoF^d^ | 0.946 | 0.931 | 0.729 | 0.709 | 0.868 | 0.918 |
|  |  |  | Missense | 0.364 | 0.368 | 0.309 | 0.286 | 0.456 | 0.646 |

^a^FNSS=familial natural short sleep; ^b^DSP=delayed sleep phase disorder; ^c^FASP=familial advanced sleeo phase; ^d^LoF=loss-of-function; ^e^Sleep duration analysed on original unit scale; ^f^Sleep duration inverse-normalised prior to analysis.

Supplementary Table 14. P-values from burden testing of rare (MAF < 0.01%) loss-of-function and missense variants in genes previously reported to harbour variants causal for disruptive sleep duration or timing on accelerometer estimates of sleep timing in UK Biobank. There were no remaining loss-of-function carriers for *GRM1*, *ADRB1* and *CRY2* within the subset of individuals from UK Biobank who wore an accelerometer.

| **Gene** | **Canonical**  **Transcript** | **Reported**  **Trait** | **Variant**  **Class** | **Sleep-midpoint^e^** | **Sleep-midpoint^f^** | **L5 Timing^g^** | **L5 Timing^h^** |
| --- | --- | --- | --- | --- | --- | --- | --- |
| *GRM1* | ENST00000361719 | FNSS^a^ | LoF^d^ | NA | NA | NA | NA |
|  |  |  | Missense | 0.910 | 0.944 | 0.277 | 0.287 |
| *NPSR1* | ENST00000359791 | FNSS^a^ | LoF^d^ | 0.586 | 0.601 | 0.682 | 0.698 |
|  |  |  | Missense | 0.968 | 0.993 | 0.480 | 0.485 |
| *ADRB1* | ENST00000369295 | FNSS^a^ | LoF^d^ | NA | NA | NA | NA |
|  |  |  | Missense | 0.015 | 0.021 | 0.945 | 0.958 |
| *BHLHE41* | ENST00000242728 | FNSS^a^ | LoF^d^ | 0.476 | 0.452 | 0.529 | 0.572 |
|  |  |  | Missense | 0.799 | 0.818 | 0.326 | 0.362 |
| *CRY1* | ENST00000008527 | DSPD^b^ | LoF^d^ | 0.508 | 0.544 | 0.993 | 0.970 |
|  |  |  | Missense | 0.011 | 0.009 | 0.238 | 0.233 |
| *PER3* | ENST00000361923 | FASP^c^ | LoF^d^ | 0.011 | 0.016 | 3.8E-06 | 8.6E-06 |
|  |  |  | Missense | 0.722 | 0.776 | 0.880 | 0.903 |
| *PER2* | ENST00000254657 | FASP^c^ | LoF^d^ | 5.1E-07 | 1.5E-06 | 9.2E-04 | 7.3E-04 |
|  |  |  | Missense | 0.500 | 0.551 | 0.261 | 0.277 |
| *CRY2* | ENST00000443527 | FASP^c^ | LoF^d^ | NA | NA | NA | NA |
|  |  |  | Missense | 0.382 | 0.343 | 0.729 | 0.770 |
| *TIMELESS* | ENST00000553532 | FASP^c^ | LoF^d^ | 0.143 | 0.137 | 0.016 | 0.013 |
|  |  |  | Missense | 0.971 | 0.994 | 0.988 | 0.959 |
| *CSNK1D* | ENST00000314028 | FASP^c^ | LoF^d^ | 0.767 | 0.741 | 0.397 | 0.374 |
|  |  |  | Missense | 0.623 | 0.709 | 0.735 | 0.830 |

^a^FNSS=familial natural short sleep; ^b^DSP=delayed sleep phase disorder; ^c^FASP=familial advanced sleeo phase; ^d^LoF=loss-of-function; ^e^Sleep-midpoint analysed on original unit scale; ^f^Sleep-midpont inverse-normalised prior to analysis; ^g^L5 timing analysed on original unit scale; ^h^L5-timing inverse-normalised prior to analysis.

### Supplementary Table 15. Summary of chronotype by PER2 loss-of-function carrier status in the UK Biobank

|  |  | **More or definitely a morning person** | | | **Definitely a morning person** | | | **More or definitely an evening person** | | | **Definitely an evening person** | | |
| --- | --- | --- | --- | --- | --- | --- | --- | --- | --- | --- | --- | --- | --- |
| **LoF Status** | **N** | **Controls (%)** | **Cases (%)** | **P^a^** | **Controls (%)** | **Cases (%)** | **P^a^** | **Controls (%)** | **Cases (%)** | **P^a^** | **Controls (%)** | **Cases (%)** | **P^a^** |
| Non-Carrier | 170,015 | 74,360 (43.7) | 95,655 (56.3) | <0.0001 | 129,577 (76.2) | 40,438 (23.8) | <0.0001 | 113,103 (66.5) | 56,912 (33.5) | 0.0001 | 156,560 (92.1) | 13,455 (7.9) | 0.009 |
| Carrier | 64 | 9 (14.1) | 55 (85.9) |  | 27 (42.2) | 37 (57.8) |  | 57 (89.1) | 7 (10.9) |  | 64 (100) | 0 (0.0) |  |

^a^P-value derived from 2-sided Fisher’s Exact test.

### Supplementary Table 16. Summary of sleep-midpoint by *PER2* loss-of-function carrier status in the UK Biobank

|  | **All Nights** | | | | | **Sleep Midpoint (Weeknights)** | | | | | **Sleep Midpoint (Weekend nights)** | | | | |
| --- | --- | --- | --- | --- | --- | --- | --- | --- | --- | --- | --- | --- | --- | --- | --- |
| **Carrier Status** | **N** | **Min^a^** | **Max^b^** | **Mean (SD^c^)** | **P^d^** | **N** | **Min^a^** | **Max^b^** | **Mean (SD^c^)** | **P^d^** | **N** | **Min^a^** | **Max^b^** | **Mean (SD^c^)** | **P^d^** |
| Non-Carrier | 34,585 | 19.45 | 30.59 | 27.01 (0.85) | <0.0001 | 34,550 | 19.22 | 30.59 | 26.92 (0.91) | <0.0001 | 33,677 | 18.92 | 32.68 | 27.27 (1.26) | 0.029 |
| Carrier | 16 | 24.04 | 27.11 | 26.05 (0.99) |  | 16 | 24.04 | 27.23 | 25.9 (1.06) |  | 15 | 25.04 | 28.21 | 26.56 (0.97) |  |

^a^Minimum; ^b^Maximum; ^c^Standard Deviation; ^d^P-value derived from 2-sided t-test.

### Supplementary Table 17. Summary of L5-midpoint timing by *PER2* loss-of-function carrier status in the UK Biobank

|  | **All nights** | | | | | **Weeknights** | | | | | **Weekend nights** | | | | |
| --- | --- | --- | --- | --- | --- | --- | --- | --- | --- | --- | --- | --- | --- | --- | --- |
| **Carrier Status** | **N** | **Min^a^** | **Max^b^** | **Mean (SD^c^)** | **P^d^** | **N** | **Min^a^** | **Max^b^** | **Mean (SD^c^)** | **P^d^** | **N** | **Min^a^** | **Max^b^** | **Mean (SD^c^)** | **P^d^** |
| Non-Carrier | 34,748 | 21.34 | 31.51 | 27.32 (1) | 0.027 | 34,713 | 20.09 | 31.80 | 27.29 (1.05) | 0.022 | 33,358 | 12.15 | 35.91 | 27.41 (1.43) | 0.458 |
| Carrier | 16 | 24.71 | 27.88 | 26.76 (0.85) |  | 16 | 24.71 | 28.24 | 26.69 (0.94) |  | 14 | 26.05 | 28.30 | 27.13 (0.63) |  |

^a^Minimum; ^b^Maximum; ^c^Standard Deviation; ^d^P-value from 2-sided t-test.
