## Supplementary Figures for "The impact of Mendelian sleep and circadian genetic variants in a population setting"

**Supplementary Figure 1**. Accelerometer traces for the four homozygous carriers in UK Biobank of the c.1657+3A>C variant in *CRY1* (A-D) as output by the GGIR R Package.

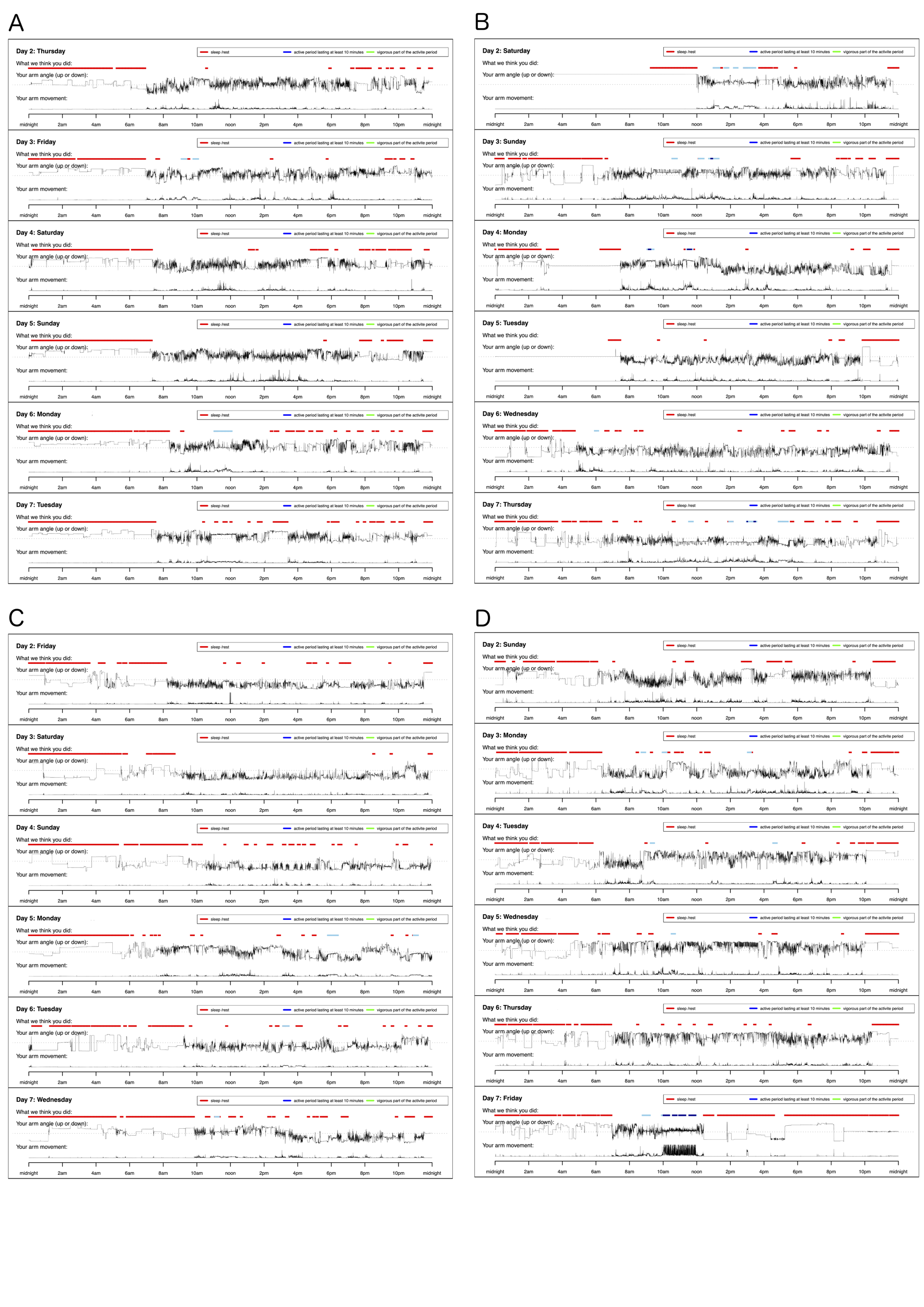

**C**

**Supplementary Figure 2**. Accelerometer traces for four heterozygous carriers in UK Biobank of the c.1657+3A>C variant in *CRY1* (A-D) as output by the GGIR R Package. Accelerometer traces shown chosen at random.

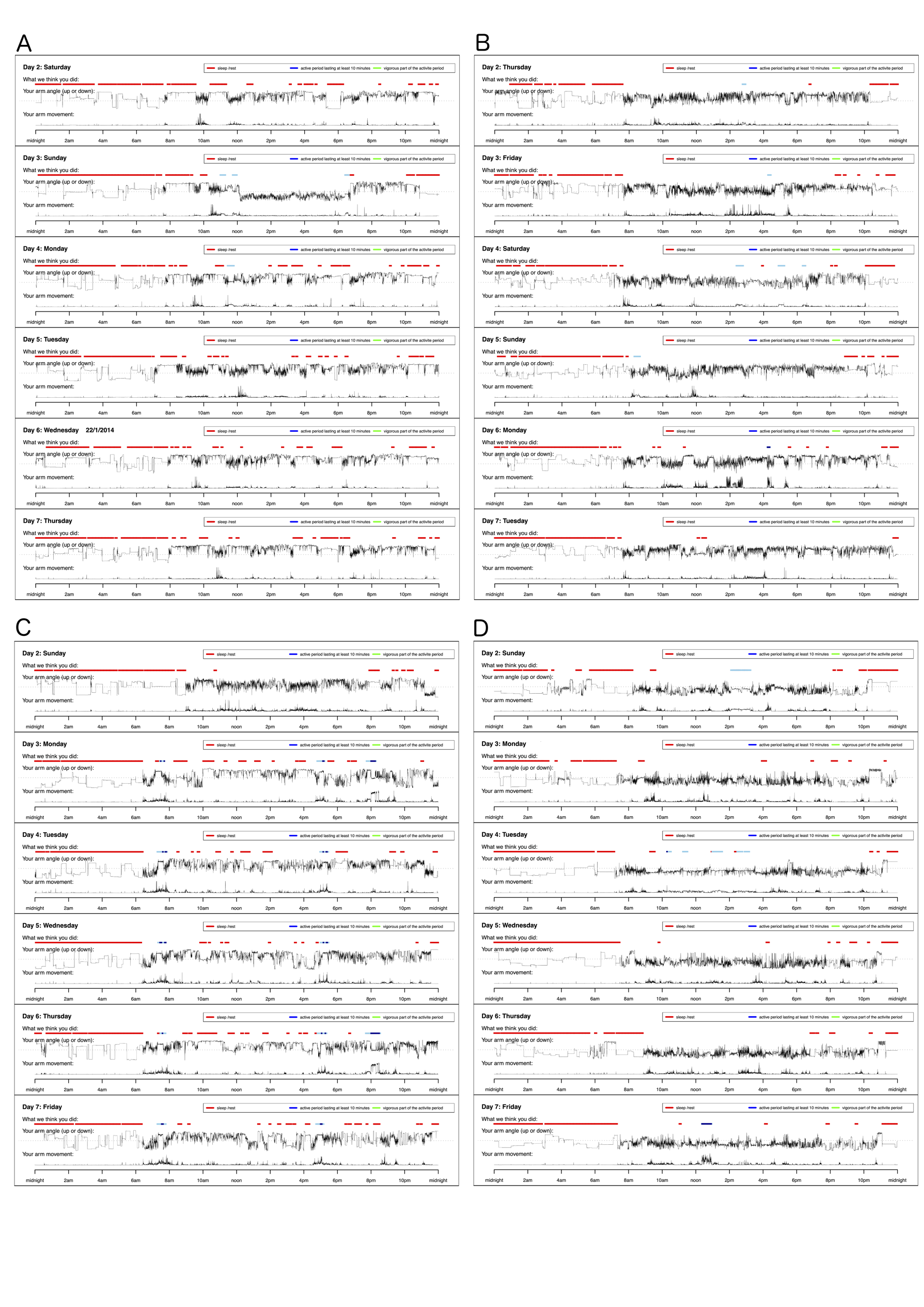

**Supplementary Figure 3**. Accelerometer traces for four individuals in UK Biobank who do not carry the c.1657+3A>C variant in *CRY1* (A-D) as output by the GGIR R Package. Accelerometer traces shown chosen at random.

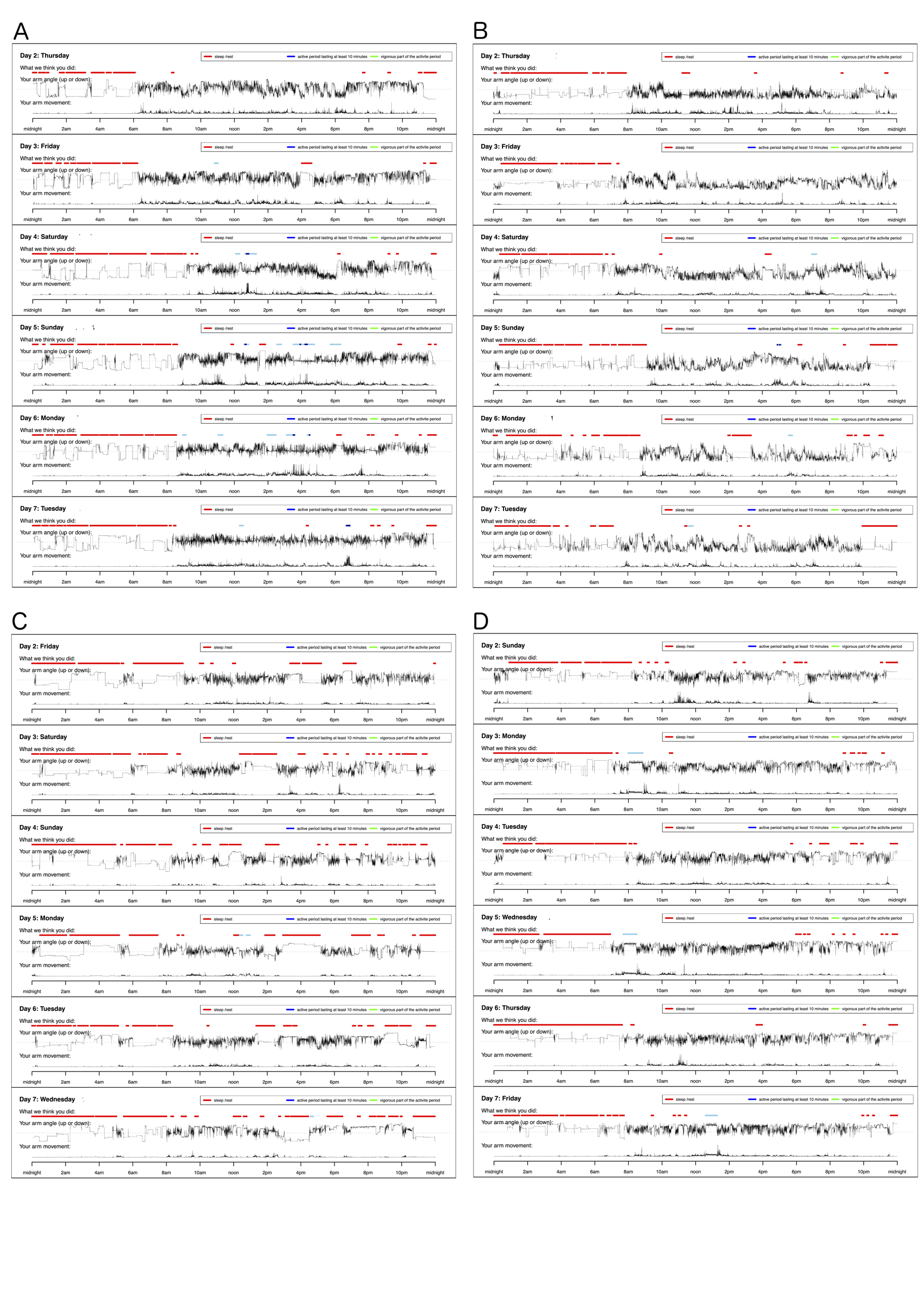
